# Examining the association between genetic risk for depression, wellbeing and schizophrenia, and proximity to greenspace

**DOI:** 10.1101/2022.04.21.22274122

**Authors:** Zoe E. Reed, Tim T. Morris, Oliver S.P. Davis, George Davey Smith, Marcus R. Munafò, Gareth J. Griffith

## Abstract

Previous studies indicate that residing in areas with greater greenspace is associated with better mental health and wellbeing. It is unclear whether these associations reflect those with better mental health seeking out greener environments.

To examine this we 1) test associations between depression, wellbeing and schizophrenia polygenic scores (PGS) with two greenspace measures in UK Biobank (N=238,306 and 293,922), 2) estimate multilevel-models (MLM), clustering individuals by local geography to investigate whether observed global effect estimates may be capturing between area differences and, 3) conduct one-sample Mendelian randomisation (MR) to estimate causal effects.

Depression and schizophrenia PGS were associated with residing in areas with lower greenspace, whilst wellbeing PGS was associated with higher greenspace. Locally-clustered MLM demonstrated attenuation for the individual wellbeing PGS association and a reversal of effect for the schizophrenia PGS association. MR revealed evidence of a causal effect of increased depression liability on decreased greenspace.

Our study provides evidence for a putative causal relationship whereby people with greater liability to depression may select into less green neighbourhoods. Our results also highlight the potential for apparently individual-level PGS effect estimates to be biased by contextual, between-area differences in outcome, which are not effectively addressed using traditional principal component adjustment.

## Introduction

Living in areas with greater greenspace has been shown to associate with both decreased prevalence (and symptoms) of mental health conditions and increased wellbeing (Gong et al., 2016; Zhang et al., 2020). Evidence has been found across many indicators of greenspace, measured in terms of both land use (Feng & Astell- Burt, 2017; White et al., 2013) and vegetation (Bezold et al., 2018a; Herrera et al., 2018; Wang et al., 2019), and across mental health outcomes, for instance, decreased psychological distress and depression (Bezold et al., 2018a, 2018b; Li et al., 2018; Sarkar et al., 2018) and increased wellbeing (Ward et al., 2016). In this research we consider two of the most commonly used and widely validated indicators. The first uses the natural reflective properties of photosynthesising plants to infer vegetation density from satellite imagery (Rhew et al., 2011), and the second uses administrative data from the land use database to estimate greenspace availability (Department for Communities and Local Government, 2007).

Similarly, residing in more urban areas, which have less greenspace, is associated with increased prevalence of mental health conditions (Faris & Dunham, 1939), including depression and schizophrenia (Pedersen, 2015; Peen et al., 2010; Purtle et al., 2019; Solmi et al., 2017; Vassos et al., 2016a). Recent meta-analyses of urbanicity and mental health studies found that risk for a range of mental health conditions was higher in urban areas (Peen et al., 2010; Purtle et al., 2019), with odds ratios (ORs) between 1.13 and 1.33. A recent Danish population-based study found incidence rate ratios between 1.15 and 2.17 for the most urban areas compared to rural areas across multiple mental health conditions (Vassos et al., 2016b). Greenspace has been suggested to drive a component of urban-rural differences in health outcomes (Verheij et al., 2008).

These associations are commonly ascribed to the “social drift” hypothesis (Sariaslan et al., 2016). Social drift proposes that those with greater risk of current or future mental health conditions may migrate, or “drift”, to more deprived, urban areas, driving the observed concentration of such conditions in urban centres (Faris & Dunham, 1939), and in turn be less likely to have access to greenspace (Pedersen, 2015). There is, however, a long history of research contesting the causal directionality of this relationship (Giggs, 1973, 1975). In the contrary hypothesis, socioeconomic position (SEP) or greenspace are considered causal risk factors for developing a mental health condition, rather than simply confounding the relationship between poor mental health and selective migration. Such debates are further complicated by spatial confounding, as urbanicity and greenspace are intimately related to many socio-demographic variables known to associate with mental health (Gong et al., 2016; March et al., 2008; Verheij et al., 2008). Therefore, identifying a causal effect of greenspace over that which might be caused by correlated, contextual confounds is difficult. The resulting identification issues contribute to the ongoing uncertainty around the causal nature of the mental health and greenspace, or urbanicity, relationship.

Previous studies exploring causal relationships between mental health and urbanicity found evidence for a causal effect of increased genetic risk for schizophrenia on living in more densely populated and deprived areas (Colodro-Conde et al., 2018; Sariaslan et al., 2016). Recent systematic reviews found evidence for mental health effects of environmental interventions to increase greenspace was absent for adults (Moore et al., 2018). However, risk-of-bias assessments in this study indicated that only four of the 14 studies provided robust data, so studies with more robust designs are needed to draw reliable conclusions.

Further evidence is therefore required to establish whether a causal relationship between greenspace and mental health liability does exist. Furthermore, such analyses must determine whether it is greenspace specifically and not urbanicity or SEP confounds driving these associations.

In this study, we assessed the association between genetic risk for several mental health outcomes and current residential greenspace for participants in UK Biobank. We also tested whether observed associations may be causal using one-sample Mendelian randomisation (MR) (Davey Smith & Ebrahim, 2003; Davey Smith & Hemani, 2014; Lawlor et al., 2008). MR exploits the randomisation of genotypes at conception to estimate causal effects of modifiable exposures on outcomes, here mental health on greenspace. We used polygenic scores (PGS) for depression, wellbeing and schizophrenia. We suggest that an association would support that those with a genetic predisposition for more positive mental health outcomes may preferentially select into environments with more greenspace.

## Results

### Sample characteristics

Sample characteristics for our UK Biobank sample are presented in Table 1.

**Table 1.** Sample characteristics

|  | **N with data available** | **Mean (SD) or percentage (%)** |
| --- | --- | --- |
| **Age** | 336,997 | 56.87 (8.00) |
| **Sex** | 336,997 | 53.79% females |
| **NDVI** | 238,306 | 0.16 (0.17) |
| **Percentage greenspace** | 293,922 | 36.86 (23.64) |
| **Depression diagnosis** | 336,997 | 12.53% |
| **MHQ depression** | 79,946 | 26.82% |
| **Wellbeing** | 109,943 | 4.59 (0.78) |
| **Schizophrenia diagnosis** | 293,922 | 0.17% |

*SD=Standard Deviation, NDVI=Normalised Difference Vegetation Index, MHQ=Mental Health*
*Questionnaire*

### Phenotypic associations

Results from linear regression models between depression, wellbeing and schizophrenia and two greenspace outcomes, adjusting for age and sex are presented in Supplementary Table S1. The two measures of greenspace we used were the mean Normalised Difference Vegetation Index (NDVI) and the percentage of an area within a 300m radius of the participants home classified as greenspace (see Methods for further details). We found negative associations between the depression and schizophrenia diagnosis phenotypes and greenspace measures, although there was weaker evidence of this for the Mental Health Questionnaire (MHQ) depression measure. We found positive associations between wellbeing and the greenspace measures, although evidence for this was weaker with the NDVI outcome.

### Linear regression model associations between polygenic scores and NDVI and percentage greenspace

Results from the linear regression model adjusting for age, sex and the first 25 principal components (PCs) are presented in Table 2 for NDVI and Table 3 for percentage greenspace. Table 2 and 3 present results for PGS calculated at a p-value threshold of 0.05 (see Supplementary Tables S2 and S3 for other p-value thresholds). Polygenic risk for depression was negatively associated with residing in green areas for both NDVI (*b*=-0.008; 95% CI: -0.01 to -0.004; *p*=0.0001) and percentage greenspace (*b*=-0.005; 95% CI: -0.01 to -0.001; *p*=0.01). This was in a broadly consistent direction across most PGS p-value thresholds (Supplementary Tables S2 and S3); however, we only found strong evidence of this association at less stringent thresholds (1x10^-03^ for the NDVI measure and 0.01 for percentage greenspace).

**Table 2.** Associations between polygenic scores for depression, wellbeing and schizophrenia and NDVI for p-value threshold of 0.05 (N=238,306)

| Phenotype | Beta* | 2.5% CI | 97.5% CI | P |
| --- | --- | --- | --- | --- |
| Depression | -0.008 | -0.01 | -0.004 | 0.0001 |
| Wellbeing | -0.0004 | -0.004 | 0.003 | 0.83 |
| Schizophrenia | 0.0005 | -0.004 | 0.005 | 0.80 |

CI=Confidence Interval. \*Beta reflects the NDVI SD change predicted by an SD increase in the PGS

**Table 3.**
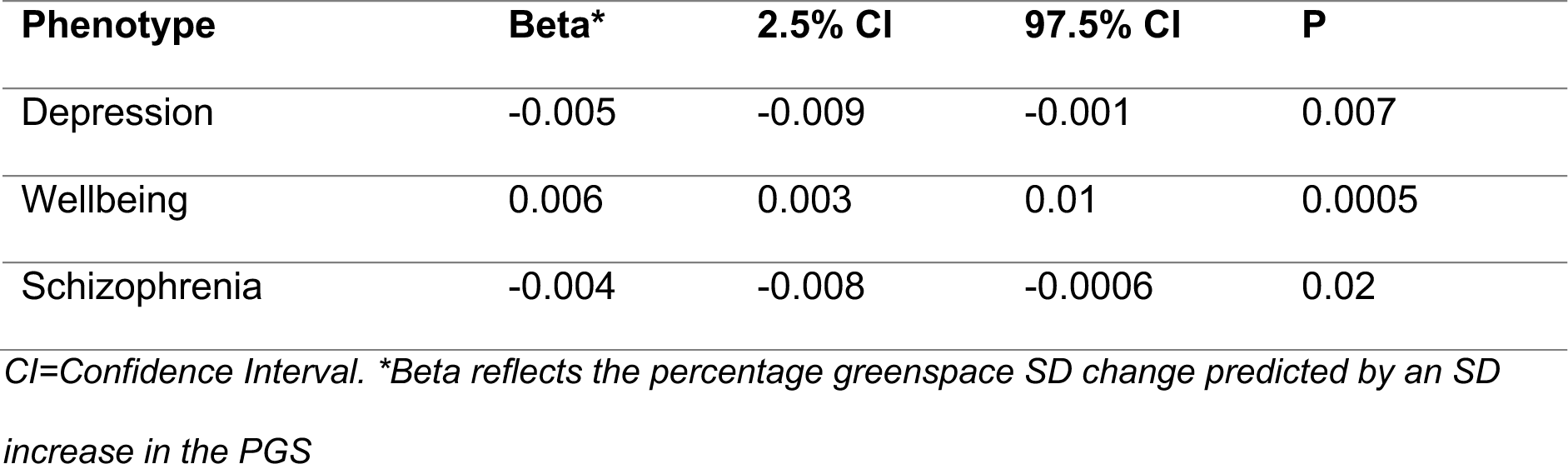
Associations between polygenic scores for depression, wellbeing and schizophrenia and percentage greenspace for p-value threshold of 0.05 (N=293,922)

The wellbeing PGS was positively associated with residing in greener areas for percentage greenspace only (*b*=0.01; 95% CI: 0.003 to 0.01; *p*=0.0005). This was in a consistent direction across all PGS p-value thresholds. However, we again only found strong evidence of this using a p-value threshold above 0.01 for percentage greenspace. We did not find strong evidence of this association with the NDVI measure (*b*=-0.0004; 95% CI: -0.004 to 0.003; *p*=0.83), although the direction of effect was positive for most other p-value thresholds.

For the schizophrenia PGS, we found strong evidence of an association with percentage greenspace only (*b*=-0.004; 95% CI: -0.01 to -0.001; *p*=0.02). This was in a consistent direction across most PGS p-value thresholds, although there is only strong evidence of this using a p-value threshold above 0.05 (Supplementary Table S3). We did not find evidence of an association with the NDVI measure (*b*=0.0005; 95% CI: -0.004 to 0.005; *p*=0.80).

### Multilevel model associations between polygenic scores and NDVI and percentage greenspace, accounting for census area

The random-intercept models allowing neighbourhood or district differentials both fit better than the linear model (Supplementary Table S4 shows comparisons of fit statistics for all three models). The level-2 VPC, which assesses the proportion of total, unexplained variation in outcome accounted for by a specific level in the model (see Supplementary Material Section 1 for further details), for the neighbourhood and district models respectively were 0.83 and 0.52 for the NDVI measure, and 0.48 and 0.24 for percentage greenspace. This implies that after adjusting for mental health PGS (across any measure), and 25 principal components, 83% of the remaining unexplained variation in individual-level NDVI exposure can be explained by the neighbourhood of the individual, or equally (from the ICC definition) we would expect a correlation of 0.83 between NDVI measures taken from pairs of individuals within the same neighbourhood.

**Table 4.** Associations between polygenic scores for depression, wellbeing and schizophrenia and NDVI measure, with neighbourhood or district level random intercepts, for p-value threshold of 0.05 (N=238,306).

| Phenotype | Neighbourhood |  |  |  | District |  |  |  |
| --- | --- | --- | --- | --- | --- | --- | --- | --- |
|  | Beta* | 2.5% CI | 97.5% CI | P | Beta* | 2.5% CI | 97.5% CI | P |
| Depression | -0.002 | -0.003 | -0.0003 | 0.02 | -0.004 | -0.006 | -0.002 | 2.13x10<sup>-04</sup> |
| Wellbeing | 0.001 | -0.0006 | 0.003 | 0.22 | 0.002 | -0.0004 | 0.004 | 0.11 |
| Schizophrenia | -0.0006 | -0.002 | 0.001 | 0.45 | -0.0007 | -0.003 | 0.001 | 0.52 |

Neighbourhood= Middle Layer Super Output Area or intermediate zone, District= Local Authority
District or Local Authority, CI=Confidence Interval. \*\*Beta reflects the NDVI SD change predicted by an SD increase in the PGS, for the average individual in a typical neighbourhood or district.

Results from these models are presented in Table 4 for NDVI (and Supplementary Table S5) and Table 5 for percentage greenspace (and Supplementary Table S6). Results from these models are comparable to those from the linear model for depression liability with NDVI under both the neighbourhood specification (*b*=-0.002; 95% CI: -0.003 to -0.0004; *p*=0.01) and the district specification (*b*=-0.004; 95% CI: - 0.01 to -0.002; *p*=2.60x10^-04^). They were also comparable for the percentage greenspace measure for neighbourhood (*b*=-0.004; 95% CI: -0.01 to -0.001; *p*=5.31x10^-03^) and district specifications (*b*=-0.01; 95% CI: -0.01 to -0.004; *p*=1.61x10^-05^). For these variables, within-area effect estimates were consistent direction with global effect estimates.

**Table 5.** Associations between polygenic scores for depression, wellbeing and schizophrenia and percentage greenspace with neighbourhood or district level random intercepts, for p-value threshold of 0.05 (N=293,922).

| Phenotype | Neighbourhood |  |  |  | District |  |  |  |
| --- | --- | --- | --- | --- | --- | --- | --- | --- |
|  | Beta* | 2.5% CI | 97.5% CI | P | Beta* | 2.5% CI | 97.5% CI | P |
| Depression | -0.004 | -0.006 | -0.001 | 6.06x10<sup>-03</sup> | -0.007 | -0.01 | -0.004 | 1.10x10<sup>-05</sup> |
| Wellbeing | 0.002 | -0.0006 | 0.005 | 0.14 | 0.004 | 0.0009 | 0.007 | 0.01 |
| Schizophrenia | 0.003 | 0.0003 | 0.006 | 0.03 | 0.001 | -0.002 | 0.004 | 0.51 |

Neighbourhood= Middle Layer Super Output Area or intermediate zone, District= Local Authority
District or Local Authority, CI=Confidence Interval. \*Beta reflects the percentage greenspace SD change predicted by an SD increase in the PGS, for the average individual in a typical neighbourhood or district.

**Table 6.** One-sample Mendelian randomisation results

| Exposure | Outcome | N | Beta* | 2.5% CI | 97.5% CI | P |
| --- | --- | --- | --- | --- | --- | --- |
| Depression<br>diagnosis | NDVI | 238,306 | -0.26 | -0.74 | 0.23 | 0.30 |
| MHQ depression | NDVI | 64,740 | -0.01 | -0.51 | 0.50 | 0.97 |
| Depression<br>diagnosis | Percentage<br>greenspace | 293,922 | -0.51 | -0.93 | -0.10 | 0.01 |
| MHQ depression | Percentage<br>greenspace | 77,358 | -0.54 | -1.14 | 0.06 | 0.08 |
| Wellbeing<br>(GW PGS) | Percentage<br>greenspace | 97,099 | 0.16 | -0.49 | 0.81 | 0.63 |
| Wellbeing<br>(PGS at $1 \times 10^{-05}$) | Percentage<br>greenspace | 97,099 | 0.06 | -0.75 | 0.87 | 0.88 |
| Schizophrenia | Percentage<br>greenspace | 293,922 | 6.75 | -7.95 | 21.45 | 0.37 |

*CI=Confidence Interval, GW=Genome-wide, PGS=Polygenic score, NDVI=Normalised Difference*
*Vegetation Index, MHQ=Mental Health Questionnaire. \*Beta reflects the NDVI or percentage*
*greenspace SD change predicted by an increased genetic liability for depression and schizophrenia*
*or every one unit increase in the wellbeing score*

The coefficient for wellbeing and percentage greenspace attenuated in both the neighbourhood specification (*b*=0.002; 95% CI: -0.001 to 0.005; *p*=0.12) and district specification (*b*=0.004; 95% CI: 0.001 to 0.01; *p*=0.01), indicating that the initial association may reflect aggregate differences in the wellbeing PGS between neighbourhoods and/or districts. However, there was still no clear evidence of an association with the NDVI measure in either specification.

Finally, for schizophrenia liability we still did not find any evidence of an effect on NDVI in either neighbourhood or district specification. However, for the percentage greenspace measure we observed an interesting effect in that the direction of the association was reversed in both the neighbourhood (*b*=0.003; 95% CI: 0.0002 to 0.01; *p*=0.03) and district specifications (*b*=0.001; 95% CI: -0.002 to 0.005; *p*=0.50). There was weaker evidence of this for the district specification; however, for the neighbourhood specification this effect was observed for all p-value thresholds. In specifying a random intercept model, we allow for between-neighbourhood (and between-district) differences in NDVI to be estimated as normally distributed residuals from a global mean. This means that the beta estimates functionally represent a within-area estimate, for instance, the effect of PGS on NDVI, given that you are in a neighbourhood with a higher-than-average NDVI. The change in direction of this sign implies that although the global relationship between schizophrenia PGS and greenspace was negative, this is mostly driven by neighbourhoods with high NDVI tending to have lower than average PGS for schizophrenia. When we account for between-area differences in our intercept, we see that within-neighbourhoods the relationship was positive. This can be considered an example of Simpson’s paradox(Hernán et al., 2011; Julious & Mullee, 1994) where an individual model would fail to address the important context of neighbourhood in determining an individual’s NDVI.

### One-sample Mendelian randomisation for depression and wellbeing on greenspace measures

We used one-sample MR analyses to assess whether there was evidence of a causal effect of depression liability on the NDVI measure and percentage greenspace, of wellbeing on percentage greenspace, and of schizophrenia liability on percentage greenspace. Results from the one-sample MR analyses are shown in Table 6 and results from sensitivity analyses are presented in Supplementary Table S7. We found strong evidence for a causal effect of the liability of having a diagnosis of depression on lower percentage greenspace (*b*=-0.51; 95% CI: -0.93 to -0.10; *p*=0.01). This was not apparent for the MHQ depression measure, although the direction of effect was the same. We did not find strong evidence of a causal effect of liability for the depression measures on the NDVI outcome; however, the direction of effect was the same for both measures. We did not find strong evidence of a causal effect of wellbeing on percentage greenspace for the PGS constructed for genome- wide significant SNPs or at the p-value threshold of 1x10^-05^, although the direction of effect was positive. Finally, we did not find strong evidence of a causal effect of schizophrenia liability on percentage greenspace.

We did not find any strong evidence for causal effects in the sensitivity analyses (Supplementary Table S7), although the direction of effect was consistent with our main results for depression liability and percentage greenspace.

## Discussion

Our results indicate that genetic risk for depression and wellbeing are associated with the greenspace of the areas which participants reside in. We found the strongest evidence for depression, with increased genetic risk associated with decreased greenspace for two different measures of greenspace. We also found evidence that this effect might be causal for percentage greenspace – in other words, that those at a higher genetic risk for depression may select into less green areas. Our MR sensitivity analyses did not provide strong evidence of a causal effect; therefore, our results should be interpreted with caution. However, the sensitivity analyses used individual SNPs and our results may therefore have been impacted by low statistical power. The positive association with wellbeing was only observed for the percentage greenspace measure, but the direction of effect was the same with the NDVI measure. Our results support previous observational studies which found an association between greenspace and depression and wellbeing (Bezold et al., 2018b, 2018a; Peen et al., 2010; Vassos et al., 2016a; Zhang et al., 2020) and extend these findings by suggesting a causal effect of liability to depression diagnosis on greenspace. As greenspace cannot cause genetic risk for these mental health outcomes, this overcomes issues around reverse causation.

In addition, models specifying a random intercept specification for local context indicated that the relationship for depression and greenspace was negative both within- and between-neighbourhoods and districts. Our concern that global effect estimates may produce misleading PGS relationships was borne out for all phenotypes, as the inclusion of an area-level random intercept attenuated results for depression liability and wellbeing, and inverted the sign of the schizophrenia liability relationship. For this study (and, we argue, for other genetically informed study designs) the effect estimate of interest is the relationship between PGS and outcome over and above that which is due to area-level compositional differences and other residual area-level confounding.

This suggests that at least part of the PGS-greenspace relationships we see were due to aggregate between-area differences, and not due to the impact of those PGS within an area. For instance, attenuation for random effects model estimates for wellbeing with percentage greenspace suggests that the initial association was likely due to individuals’ wellbeing PGS being consistent within an area. Thus, when we accounted for between-area differences via the random intercept specification, the observed relationship within-area became far less strong, as most of this initial relationship was driven by certain areas having both high wellbeing PGS and high greenspace levels.

This is evidenced more explicitly by the extremely high level-2 VPC values. We note that these were not identical for all mental health PGS but the amount of variance explained by each PGS was very small, so the VPC was almost the same for each model. A VPC of 0.83 implies that simply knowing individuals’ neighbourhoods would allow you to account for 83% of the unexplained, observed variation in phenotypic NDVI. This is important as, coupled with the reversal of sign of the effect estimate in our schizophrenia liability model, it implies our initial global effect estimate was mostly driven by individuals living within the same area (both neighbourhood or district areas) being more phenotypically and genetically similar to their immediate neighbours than to those living in other areas. This is particularly concerning as this is what adjustment for genetic principal components is often cited as insulating against, with the implication that adjustment for genetic ancestry should deal with spatial confounding between UK Biobank participants. This appears to not be the case, and as such we propose that local neighbourhood random intercept specifications be included in PGS analyses as a routine robustness check to ensure that global PGS estimates are not being biased by such spatial confounding. That said, our study design is likely an extreme case of such confounding, as the greenspace value for any given individual can be reasonably inferred to be nearly identical of that of their immediate neighbours, simply because the outcome is defined within a spatial boundary.

Our results for depression and wellbeing PGS are consistent with the social drift hypothesis, which supports a role of the movement of genetically similar individuals into more urban/less green areas. Here, this suggests those with greater genetic risk of poor mental health may select into areas with less greenspace. We did not test a bidirectional effect for mental health with greenspace as we were unable to construct strong genetic instruments for greenspace. We did run GWAS for both greenspace measures in UK Biobank, but only found one genome-wide significant SNP for NDVI, and none for percentage greenspace so analyses would be impossible, or underpowered to detect any effects. If larger GWAS of contextual phenotypes were to be conducted, this may help investigate causation in this direction.

The relationship we observe for schizophrenia liability is particularly interesting. We found that for both greenspace measures when we modelled the association with schizophrenia liability the direction of the association reversed when specifying an area-level random-intercept parameter, an example of Simpson’s paradox(Kievit et al., 2013). There are multiple reasons why this may occur, but one which is consistent with known UK Biobank selection issues is that this is caused by non-random selection within areas. UK Biobank participation is known to be patterned by both phenotypic schizophrenia and schizophrenia PGS(Tyrrell et al., 2021). We suggest that the results we see are consistent with collider bias where, within-areas, those with low schizophrenia PGS and in greener areas are more likely to have participated in UK Biobank. Assuming no complex interactions, this would induce a positive relationship between schizophrenia PGS and greenness as a result of collider bias, where individuals with a high schizophrenia PGS are more likely to have high greenspace values. What is striking, however, is that schizophrenia PGS and greenspace are still clustered strongly enough that between areas we see the anticipated relationship, implying that areas with higher greenspace tend to be populated with individuals with lower schizophrenia PGS to a strong enough degree that the global effect estimate is negative in the full UK Biobank population.

Our results also differed between the NDVI and greenspace measure used. Our results are consistent with previous observational studies showing increased NDVI, which indicates visible greenness of an area was associated with fewer depressive symptoms, decreased job-related stress and psychological distress (Bezold et al., 2018a; Herrera et al., 2018; Wang et al., 2019). However, in other studies using a similar measure to ours based on data from the Generalized Land Use Database (i.e., percentage of greenspace in an area), associations with mental health outcomes were not generally found (Feng & Astell-Burt, 2017; Mueller et al., 2019; Weeland et al., 2019).

## Limitations

Our study has several limitations. First, PGS analyses have well known limitations; for example, different p-value thresholds to construct scores affects the predictive accuracy of the score. More SNPs may increase the variance explained but may also suffer from overfitting. In our main analyses we focused on one p-value threshold but included other thresholds as sensitivity analyses (see Supplementary Tables S2 and S3) and found consistent results across these. Second, our large sample size means that the estimated associations will have increased precision and we may overstate confidence in associations which may not generalise to a wider population. Similarly, these results were obtained from a European ancestry sample and may not be generalisable to other ancestry groups. Third, it is possible that the GWAS for our mental health phenotypes also include some SNPs associated with “upstream” traits like greenspace (i.e., the primary phenotype is mis-specified) (Richmond & Davey Smith, 2021). As we do not know what SNPs influence greenspace, we are unable to account for this. However, the PRS analyses are more likely to be impacted by this than the MR analyses given the less stringent p-values used and therefore the greater number of SNPs included that may capture these “upstream” traits. Fourth, whilst we may increase the likelihood of obtaining generalisable estimates by including a random intercept for area-context, participation in UK Biobank at baseline was extremely non-random as we outline above. Fifth, we were not able to be prescriptive with our operationalisation of greenspace. A recent study illustrated the sensitivity of results to greenspace specification; for instance, availability, accessibility, and visibility of greenspace produced very different results when included in analyses (Labib et al., 2021).The NDVI measure likely offers a more bespoke measure of greenspace in rural settings as it is calculated at a household level, rather than at OA level (rural OAs are sparser and larger). However, NDVI simply captures surface reflectivity from satellite imagery, so has no sensitivity to the accessibility of the greenspace it is capturing, unlike the percentage greenspace measure. Future research into this must carefully consider which components of greenspace are of interest for each research question. Sixth, in our MR analyses we were possibly underpowered to detect an effect for wellbeing due to the small number of genome-wide significant SNPs identified in the GWAS. Seventh, a key limitation of MLM approaches is that endogenous, omitted variables associated with the higher-level contexts can reduce generalisability. If the aim of an analysis is solely to adjust for higher level context, then a fixed effects approach might be more appropriate (Abdellaoui et al., 2021).

Finally, we also need to consider that geographical location will also be, to some extent, accounted for in our PGS (Haworth et al., 2019), as well as other types of structure, which may include population stratification and assortative mating (Morris et al., 2020).

In conclusion, we found evidence to suggest that those at risk of poorer mental health outcomes, specifically depression and wellbeing, may select into less green areas. Further studies investigating bidirectionality are called for as if lack of greenspace further exacerbates poor mental health then interventions targeting greenspace may be important. Finally, our study highlights the potential for spatial confounding to bias genetic analyses of geographically clustered phenomena. We suggest therefore, that increasing availability of genetic datasets with geographical location will allow for future research to understand and quantify these potential biases.

## Methods

### Cohort description

The UK Biobank is a large population-based prospective health research resource of 503,317 participants, aged between 38 and 73 years at recruitment, recruited between 2006 and 2010, from across the UK (Sudlow et al., 2015). The study aimed to identify determinants of human disease in middle and old age. A range of data have been collected across sociodemographic, health status, lifestyle, cognitive function, self-reported measures and physical and mental health measures. Data were collected via a number of methods, including paper and web-based questionnaires, computer assisted interviews, clinic visits and data linkage. Baseline assessment took place at 22 assessment centres in the UK to enable recruitment from a range of locations. Further information can be found on the UK Biobank website (www.ukbiobank.ac.uk). UK Biobank received ethical approval from the Research Ethics Committee (REC reference for UK Biobank is 11/NW/0382). We excluded participants based on the latest withdrawal lists at the time of data extraction (04/02/2020) for our UK Biobank project (project number: 21829).

### Phenotypic measures

The proposed mechanism of greenspace impact is inconsistently defined in the literature (Cusack et al., 2017). Therefore, we used two measures to capture participants’ residential access to greenspace. These measures were made available to UK Biobank by Sarkar et al., and were derived using exact postcodes (Sarkar et al., 2015). Both measures were standardised prior to inclusion in models.

### Normalised Difference Vegetation Index

Our first outcome was the mean Normalised Difference Vegetation Index (NDVI), calculated for a 500m radius of the participant’s home location. NDVI is a commonly used parameterisation of green-space access as it can be calculated from satellite imagery, rather than requiring administrative land use information (Herrera et al., 2018; Wang et al., 2019), and closely approximates subjective exposure estimates (Rhew et al., 2011). The NDVI ranges between -1 and +1, with positive values indicating greener areas based on reflectance measures in Colour InfraRed (CIR) satellite data(Sarkar et al., 2015). The distribution in UK Biobank was -0.50 to 0.70. After exclusions based on withdrawn consent and quality control filtering for genetic data (described below) and including only those with genetic data, the final sample size for NDVI was 238,306 (see Figure 1).

**Figure 1.**
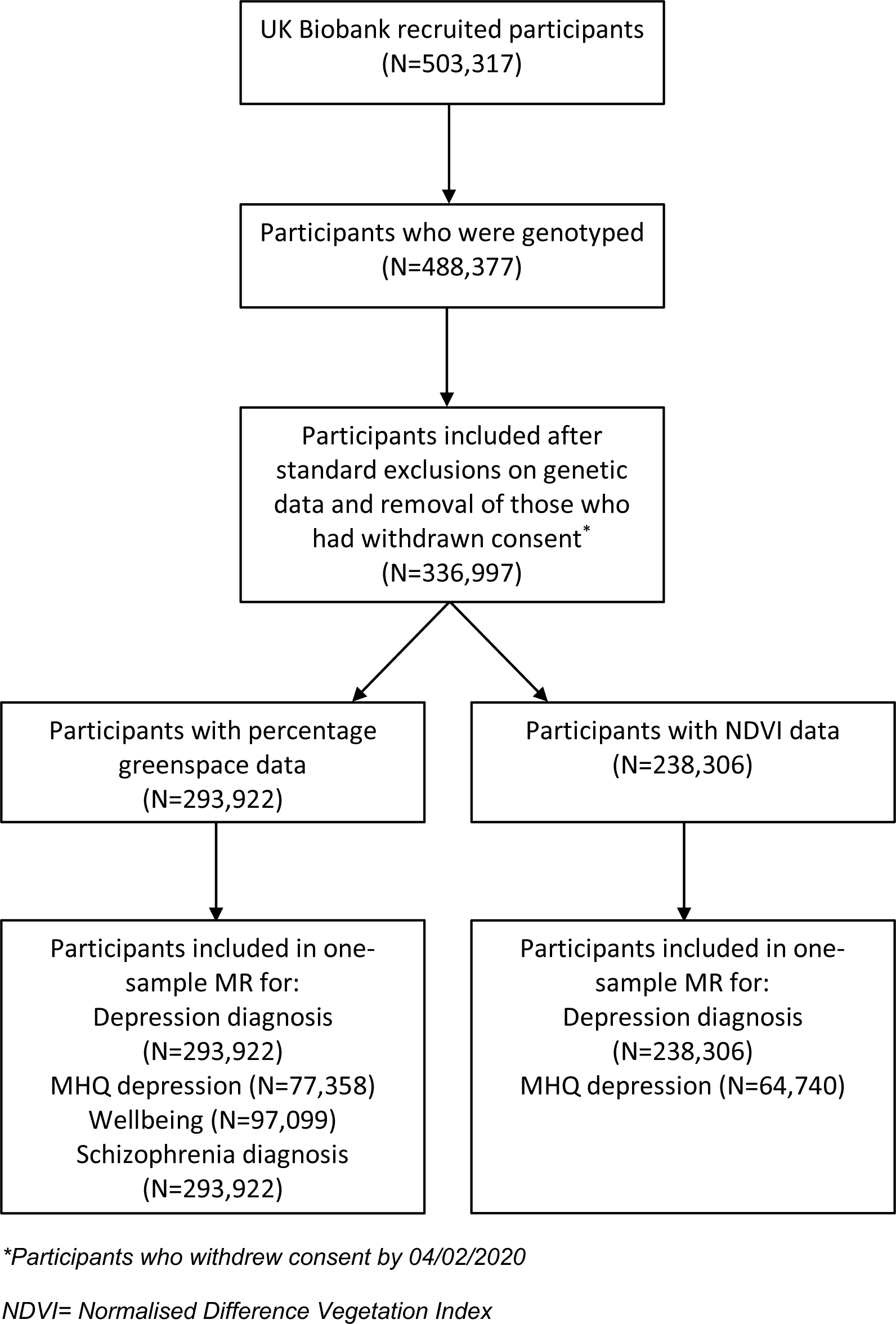
Flowchart of number of participants included in analyses

### Percentage greenspace

Our second outcome refers to the percentage of an area within a 300m radius of the participants home classified as greenspace. This is based on land use data from the Generalized Land Use Database (GLUD) (Department for Communities and Local Government, 2007) for England 2005 data at the 2001 census output area (OA) level. Census output areas were designed to have a consistent population, thus their area varies considerably with population density. Each participant’s home location was allocated an area weighted mean of these data for output areas intersecting the 300m radius of a home. After exclusions and including only those with genetic data, the final sample size used in analyses with this outcome was 293,922 (see Figure 1).

The percentage greenspace measure was only weakly correlated with the NDVI measure (*r*=0.16). This is likely because they capture different aspects of greenspace; the NDVI measure captures greenness via the reflectance of the land surface, whereas the percentage greenspace classification captures functional greenspace, including areas of natural agricultural land, recreational areas and other areas of grassland, and excluding domestic gardens (Department for Communities and Local Government, 2007).

### Depression

For one-sample MR analyses, we also require phenotypic data on mental health outcomes. We used data from a combination of both ICD-10 diagnoses and diagnoses reported in verbal interviews with a nurse at the assessment centre. Anyone who had a diagnosis of depression from any of these sources was coded as having depression. Anyone without a diagnosis indicated in these data sources was coded as not having depression. We refer to this combined measure as diagnosis of depression. Data were also available for a sub-sample of UK Biobank participants who completed questions related to depressive and manic symptoms during the final two years of recruitment. From these data, a variable for having bipolar disorder and major depression was derived, where a participant reporting both was coded only as having the most severe condition (Smith et al., 2013). We excluded participants who had bipolar disorder type 1 or 2 and identified those as having depression if they had probable recurrent major depression (severe or moderate) or a single probable major depression episode. We refer to this as the mental health questionnaire (MHQ) depression measure.

### Wellbeing

As part of an on-line mental health self-assessment questionnaire issued in 2016, participants were asked the question ‘In general how happy are you?’. Participants could select from the following options: ‘Prefer not to answer’, ‘Do not know’, ‘Extremely happy’, ‘Very happy’, ‘Moderately happy’, ‘Moderately unhappy, ‘Very unhappy’ and ‘Extremely unhappy’. We excluded participants who selected either of the first two options and coded the other options so that a higher number indicated a more positive response.

### Schizophrenia

Similar to the depression diagnosis variable, we used data from a combination of both ICD-10 diagnoses and diagnoses reported in verbal interviews with a nurse at the assessment centre. Participants with a diagnosis of schizophrenia from any of these sources were coded as having schizophrenia. Participants without a diagnosis indicated in these data sources were coded as not having schizophrenia. In UK Biobank 0.17% of participants were coded as having schizophrenia, compared to 0.7% reported for psychotic disorders in England (McManus et al., 2016). This is consistent with known selection pressures in UK Biobank , as demonstrated previously with reduced participation in individuals with greater liability to schizophrenia (Tyrrell et al., 2021).

### Potential confounders

We included age at time of initial assessment and sex in all of our analyses. As we examined phenotypes related to geographical location, we also included the first 25 principal components of population structure (PCs) as confounders in all of our models including PGS. The number of PCs was chosen to reduce geographical confounding. A recent study found that adjusting for 18 PCs was a sensible correction for population structure (Sarmanova et al., 2020), but in this study we corrected for 25 PCs as this was where we saw a flattening in the variance explained by the PCs in our outcomes (Supplementary Figure S1).

### Structural Variables

In our multilevel models we specified geographical clustering using the administrative area that each participant resided in at the time of recruitment.

Participant locations were matched on easting and northing coordinates (Office for National Statistics, 2020) to the most recently defined local administrative units at two spatial scales, one to represent local neighbourhood context, and one the broader city and commuting context. We consider Middle Layer Super Output Areas (MSOAs, average population = 7,200) for England and Wales analogous to Intermediate Zones (IZ, average population = 4,200) for Scotland – hereafter, “neighbourhoods”. Similarly, we consider Local Authority Districts (LADs, average population 140,000) analogous to Local Authorities (LAs, average population 170,000) for Scotland, hereafter “districts”. Supplementary Figures S2 and S3 show the number of UK Biobank participants per neighbourhood and district respectively. Data were rounded to the nearest kilometre to preserve anonymity, meaning some participants’ recorded locations fell outside of census boundaries (0.6% of participants). These participants were assigned to the nearest neighbourhood or district based on Euclidean distance to the census output area centre.

### Genetic data

There were 488,377 participants with genotyped samples, of which 49,979 were genotyped using the UK BiLEVE array and 438,398 using the UK Biobank axiom array. Pre-imputation quality control, phasing and imputation have been described elsewhere (Bycroft et al., 2018). In summary, multiallelic SNPs and those with a MAF≤1% were removed. Phasing of genotype data was performed using a modified version of the SHAPEIT2 algorithm. The SNPs used were imputed to the Haplotype Reference Consortium (HRC) reference panel, using IMPUTE2 algorithms. A graded filtering with different imputation qualities for different MAF ranges was used (Info>0.3 for MAF>3%, Info>0.6 for MAF 1-3%, Info>0.8 for MAF 0.5-1% and Info>0.9 for MAF 0.1-0.5%), where MAF and info scores were recalculated on an in-house derived ‘European’ subset. Individuals with sex-mismatch or sex-chromosome aneuploidy were excluded (N=814). In-house quality control filtering of the UK Biobank data is described in a published protocol(Mitchell et al., 2019). We restricted the sample to individuals of white British ancestry who self-report as “White British” and who have very similar ancestral backgrounds according to principal components analysis (N=409,703), as described by Bycroft and colleagues (Bycroft et al., 2018). Estimated kinship coefficients using the KING toolset (Manichaikul et al., 2010) identified 107,162 pairs of related individuals. An in-house algorithm was then applied to this list and preferentially removed the individuals related to the greatest number of other individuals until no related pairs remain. These individuals were excluded (N=79,450).

### Polygenic scores

We constructed 12 different PGS using Plink (version 2) (Purcell et al., 2007) for each phenotype using summary statistics from genome wide association studies (GWAS) of major depressive disorder (MDD) (Wray et al., 2018), wellbeing (Okbay et al., 2016) and schizophrenia (Ripke et al., 2014). The summary statistics for the MDD and wellbeing GWAS excluded UK Biobank to avoid sample overlap, and also excluded 23andMe participants. The PGS we constructed were based on different p-value thresholds of the trait GWAS (0.5, 0.4, 0.3, 0.2, 0.1, 0.05, 0.01, 1x10^-3^, 1x10^-4^, 1x10^-5^, 1x10^-6^, 5x10^-8^). We used the PGS constructed at a p-value threshold of 0.05 for our main analyses due to this explaining the largest proportion of variance in depression (Wray et al., 2018) and schizophrenia (Ripke et al., 2014) in the original GWAS studies. This was not reported for wellbeing; therefore, we used a p-value threshold of 0.05 for these analyses for consistency. We present results from other p-value thresholds as sensitivity analyses to assess the consistency of the observed effects across different thresholds. For the PGS constructed at a p-value threshold of 0.05, the depression PGS explained 0.4% of the variance in having a diagnosis of depression in the UK Biobank, the wellbeing PGS explained 0.3% of the variance in wellbeing, and the schizophrenia PGS explained 2.7% of the variance in having a diagnosis of schizophrenia. SNPs from the GWAS and UK Biobank were harmonised, aligning the effect estimates and alleles. SNPs were clumped used the European subsample of the 1000 genomes project, with R^2^ < 0.25 and a window of 500 kb. The PGS were created by multiplying the number of effect alleles for each participant in UK Biobank by the effect estimate of the SNP from the relevant GWAS, then summing across all SNPs associated with each trait. PGS were z-standardised; results should be interpreted as per standard deviation (SD) increase in score.

### Statistical analysis

#### Linear regression models

Initial analyses were conducted in R version 3.5.1 and 3.6.2 (R Core Team, 2016) using linear regression models. We first conducted analyses to examine phenotypic associations in UK Biobank where NDVI or percentage greenspace were the outcomes and the mental health phenotypes were the exposures. We then conducted analyses where NDVI or percentage greenspace were the outcomes and the PGS for MDD, wellbeing and schizophrenia were the exposures in separate analyses. These global regression models cannot meaningfully distinguish within- and between-area components of a given relationship but are useful to demonstrate aggregate relationships across a dataset. It also allows us to observe whether there are differences when accounting for geographical context which may indicate contextually varying relationships.

#### Multilevel models (MLM)

Our interest for this study is in modelling the impact of the mental health PGS on greenspace, but also in quantifying the degree to which the ostensibly individual relationship can be distorted by higher level spatial confounding. As such, we fit multilevel (or mixed or hierarchical) models (MLM) to efficiently estimate the within-area effect of PGS exposure, whilst we account for between-area differences in greenspace and PGS, retaining the capacity to quantify the impact of contextual clustering that is sacrificed in traditional fixed effect approaches (Bell & Jones, 2015). We present this as a Variance Partitioning Coefficient (VPC), for the area level, which gives the proportion of unexplained variation in a coefficient, that is explained by the area level rather than solely being assumed a result of individual differences. In the two level case, this is exactly equivalent to the intra-class correlation (ICC), giving the expected correlation between randomly selected pairs of individuals within the same higher level unit (Merlo et al., 2005). This ensures we do not misattribute between-area aggregate variation in mental-health PGS exposures, to independent between-individual variation. The MLM’s are further described in the Supplementary Material (Section 1).

For MLMs we used the R package *lme4* (Bates et al., 2015). We fitted two MLMs, adjusting for sex, age and 25 PCs as fixed effects. The first model included a normally distributed random effect around the modelled intercept, which varies at neighbourhood level. By allowing each area to have its own modelled intercept, we account for the fact that different geographical areas have different mean values for exposure and outcome. This means our PGS effect estimate represents the effect of a unit increase in PGS *within* a given neighbourhood. The second MLM extends the formulation above but with random effects specified for district rather than neighbourhood. We evaluated the model fit using Akaike’s Information Criteria (AIC) and compared across all three models, taking the best fitting model as the one with lowest AIC. We also present the proportion of the total residual variance attributable to variation between neighbourhoods or districts, rather than between-individual, within-area variation, by calculating ICCs.

#### One-sample Mendelian randomisation analyses

Where we found evidence of an association between the PGS and the greenspace outcomes, we ran one-sample MR analyses, with externally derived weights, to assess whether these relationships may be causal. MR is a causal inference method which uses genetic variants (typically SNPs), which are randomly assigned at conception, as instrumental variables (IVs) for an exposure of interest. MR is more robust to assumptions of confounding and reverse causation than conventional observational analyses. There are three underlying core assumptions for MR, which are i) that the genetic IV is robustly associated with the exposure (relevance), ii) the genetic IV is only associated with the outcome via the exposure i.e., no direct effects on the outcome (exclusion restriction), and iii) there is no confounding of the genetic IV and outcome (independence).

In these analyses, for the genetic IV for depression, wellbeing and schizophrenia, we used weighted PGS created using SNPs that reached genome-wide significance (p<5x10^-8^) in the GWAS including all samples. For schizophrenia we used the corresponding weights from the GWAS including all samples, however for depression and wellbeing we used weights from the GWAS excluding UK Biobank and 23andMe samples (Okbay et al., 2016; Ripke et al., 2014; Wray et al., 2018). For the genetic IV for wellbeing, we also used a weighted PGS created at p<1x10^-05^, due to the low number of SNPs (n=3) reaching genome-wide significance.

We used a two-stage least squares regression approach, with the Applied Econometrics with R (AER) package (v1.2-9), to estimate the causal effects of depression liability, wellbeing and schizophrenia liability on the greenspace outcomes. Two-stage least squares regression allows the IV-exposure effect to be estimated in the first step and then in the second step the effect on the outcome is estimated by regressing the outcome on the estimates from step one. For depression, we used the variable for diagnosis of depression to estimate the genetic IV and exposure relationship in our main analyses, and we used the MHQ depression measure in sensitivity analyses. For schizophrenia we used the variable for diagnosis of schizophrenia to estimate the genetic IV and exposure relationship.

We also ran sensitivity analyses for all one-sample MR analyses using individual SNP data (as these were not possible to apply to the PGS), except for the genome- wide p-value threshold wellbeing analysis as there were too few SNPs. We used the collider-correction approach (Barry et al., 2021), which applies two-sample MR methods to one-sample MR data. This approach artificially induces collider bias between the exposure and outcome and then corrects for it, in order to facilitate simple adjustment for weak instrument bias and pleiotropy. We used collider- corrected inverse-variance weighted (IVW), Simulation Extrapolation (SIMEX) adjusted MR-Egger and least absolute deviation approaches. The IVW approach constrains the regression intercept to pass through zero and assumes there is no unbalanced pleiotropic effects of SNPs. The IVW estimate ignores uncertainty in the SNP-exposure estimate by assuming that its variance is negligible (known as the NO Measurement Error [NOME] assumption). MR Egger assesses and counteracts unbalanced pleiotropy by not constraining the intercept to pass through zero. SIMEX allows for adjustment due to regression dilution bias from weak instruments. Finally, IVW and MR Egger rely on the InSIDE assumption (that the SNP-exposure association is not correlated with a path from the SNP to the outcome, independent of the exposure) and the least absolute deviation approach does not. The least absolute deviation approach is also SIMEX adjusted. We also calculated the I-squared statistic (indicative of strength of NOME violation for MR Egger) and the mean F-statistic (indicative of instrument strength) (Bowden et al., 2016). Finally, we ran MR Robust Adjusted Profile Score (RAPS) for the wellbeing MR using a p-value threshold of p<1x10^-05^. As it allows large numbers of weak instruments below the conventional GWAS threshold (5x10^-08^) to be included, accounting for weak instrument bias.

## Data availability

UK Biobank data are available through a procedure described at https://www.ukbiobank.ac.uk/enable-your-research.

## Code availability

The analysis code that forms the basis of the results presented here is available from https://github.com/ZoeReed/Greenspace_MH_PGS_analysis.

## Acknowledgements

This research has been conducted using data from UK Biobank, a major biomedical database (www.ukbiobank.ac.uk, application No 21829). We thank the participants of UK Biobank. Quality Control filtering of the UK Biobank data was conducted by R.Mitchell, G.Hemani, T.Dudding, L.Corbin, S.Harrison, L.Paternoster as described in the published protocol (doi: 10.5523/bris.1ovaau5sxunp2cv8rcy88688v).

## Author contributions

Conceptualization: ZER, MRM, GJG; Methodology: ZER, GJG, TTM; Formal Analysis: ZER; Resources: MRM, OSPD, GDS; Data Curation: ZER; Writing—Original Draft: ZER, GJG; Writing—Review and Editing: ZER, GJG, TTM, OSPD, GDS, MRM; Supervision: MRM; Project Administration: MRM; Funding Acquisition: MRM, GDS, OSPD.

## References

Abdellaoui, A., Verweij, K. J. H., & Nivard, M. G. (2021). Geographic Confounding in Genome-Wide Association Studies. BioRxiv, 2021.03.18.435971. https://doi.org/10.1101/2021.03.18.435971

Barry, C., Liu, J., Richmond, R., Rutter, M. K., Lawlor, D. A., Dudbridge, F., & Bowden, J. (2021). Exploiting collider bias to apply two-sample summary data Mendelian randomization methods to one-sample individual level data. PLOS Genetics, 17(8), e1009703. https://doi.org/10.1371/journal.pgen.1009703

Bates, D., Mächler, M., Bolker, B. M., & Walker, S. C. (2015). Fitting linear mixed- effects models using lme4. Journal of Statistical Software, 67(1). https://doi.org/10.18637/jss.v067.i01

Bell, A., & Jones, K. (2015). Explaining Fixed Effects: Random Effects Modeling of Time-Series Cross-Sectional and Panel Data. Political Science Research and Methods, 3(1), 133–153. https://doi.org/10.1017/psrm.2014.7

Bezold, C. P., Banay, R. F., Coull, B. A., Hart, J. E., James, P., Kubzansky, L. D., Missmer, S. A., & Laden, F. (2018a). The Association Between Natural Environments and Depressive Symptoms in Adolescents Living in the United States. Journal of Adolescent Health, 62(4), 488–495. https://doi.org/10.1016/j.jadohealth.2017.10.008

Bezold, C. P., Banay, R. F., Coull, B. A., Hart, J. E., James, P., Kubzansky, L. D., Missmer, S. A., & Laden, F. (2018b). The relationship between surrounding greenness in childhood and adolescence and depressive symptoms in adolescence and early adulthood. Annals of Epidemiology, 28(4), 213–219. https://doi.org/10.1016/J.ANNEPIDEM.2018.01.009

Bowden, J., Fabiola Del Greco, M., Minelli, C., Smith, G. D., Sheehan, N. A., & Thompson, J. R. (2016). Assessing the suitability of summary data for two- sample mendelian randomization analyses using MR-Egger regression: The role of the I 2 statistic. International Journal of Epidemiology, 45(6), 1961–1974. https://doi.org/10.1093/ije/dyw220

Bycroft, C., Freeman, C., Petkova, D., Band, G., Elliott, L. T., Sharp, K., Motyer, A., Vukcevic, D., Delaneau, O., O’Connell, J., Cortes, A., Welsh, S., Young, A., Effingham, M., McVean, G., Leslie, S., Allen, N., Donnelly, P., & Marchini, J. (2018). The UK Biobank resource with deep phenotyping and genomic data. Nature, 562(7726), 203–209. https://doi.org/10.1038/s41586-018-0579-z

Colodro-Conde, L., Couvy-Duchesne, B., Whitfield, J. B., Streit, F., Gordon, S., Kemper, K. E., Yengo, L., Zheng, Z., Trzaskowski, M., De Zeeuw, E. L., Nivard, M. G., Das, M., Neale, R. E., MacGregor, S., Olsen, C. M., Whiteman, D. C., Boomsma, D. I., Yang, J., Rietschel, M., … Martin, N. G. (2018). Association between population density and genetic risk for schizophrenia. JAMA Psychiatry, 75(9), 901–910. https://doi.org/10.1001/jamapsychiatry.2018.1581

Cusack, L., Larkin, A., Carozza, S. E., & Hystad, P. (2017). Associations between multiple green space measures and birth weight across two US cities. Health and Place, 47, 36–43. https://doi.org/10.1016/j.healthplace.2017.07.002

Davey Smith, G., & Ebrahim, S. (2003). “Mendelian randomization”: can genetic epidemiology contribute to understanding environmental determinants of disease? International Journal of Epidemiology, 32(1), 1–22. https://doi.org/10.1093/ije/dyg070

Davey Smith, G., & Hemani, G. (2014). Mendelian randomization: genetic anchors for causal inference in epidemiological studies. Human Molecular Genetics, 23(R1), R89–98. https://doi.org/10.1093/hmg/ddu328

Department for Communities and Local Government. (2007). Generalised Land Use Database Statistics for England 2005 (Enhanced Basemap). DCLG.

Faris, R. E. L., & Dunham, H. W. (1939). Mental disorders in urban areas: an ecological study of schizophrenia and other psychoses. In Mental disorders in urban areas: an ecological study of schizophrenia and other psychoses. Univ. Chicago Press.

Feng, X., & Astell-Burt, T. (2017). The relationship between neighbourhood green space and child mental wellbeing depends upon whom you ask: Multilevel evidence from 3083 children aged 12–13 years. In International Journal of Environmental Research and Public Health (Vol. 14, Issue 3, p. 235). Multidisciplinary Digital Publishing Institute. https://doi.org/10.3390/ijerph14030235

Giggs, J. A. (1973). The Distribution of Schizophrenics in Nottingham. Transactions of the Institute of British Geographers, 59, 55–76. https://doi.org/10.2307/621712

Giggs, J. A. (1975). The Distribution of Schizophrenics in Nottingham: A Reply. Transactions of the Institute of British Geographers, 64, 150–156. https://doi.org/10.2307/621712

Gong, Y., Palmer, S., Gallacher, J., Marsden, T., & Fone, D. (2016). A systematic review of the relationship between objective measurements of the urban environment and psychological distress. Environment International, 96, 48–57. https://doi.org/10.1016/j.envint.2016.08.019

Haworth, S., Mitchell, R., Corbin, L., Wade, K. H., Dudding, T., Budu-Aggrey, A., Carslake, D., Hemani, G., Paternoster, L., Smith, G. D., Davies, N., Lawson, D. J., & J. Timpson, N. (2019). Apparent latent structure within the UK Biobank sample has implications for epidemiological analysis. Nature Communications, 10(1), 333. https://doi.org/10.1038/s41467-018-08219-1

Hernán, M. A., Clayton, D., & Keiding, N. (2011). The simpson’s paradox unraveled. International Journal of Epidemiology, 40(3), 780–785. https://doi.org/10.1093/ije/dyr041

Herrera, R., Markevych, I., Berger, U., Genuneit, J., Gerlich, J., Nowak, D., Schlotz, W., Vogelberg, C., Von Mutius, E., Weinmayr, G., Windstetter, D., Weigl, M., Heinrich, J., & Radon, K. (2018). Greenness and job-related chronic stress in young adults: A prospective cohort study in Germany. BMJ Open, 8(6), e021599. https://doi.org/10.1136/bmjopen-2018-021599

Julious, S. A., & Mullee, M. A. (1994). Confounding and Simpson’s paradox. BMJ (Clinical Research Ed.), 309(6967), 1480–1481. https://doi.org/10.1136/bmj.309.6967.1480

Kievit, R. A., Frankenhuis, W. E., Waldorp, L. J., & Borsboom, D. (2013). Simpson’s paradox in psychological science: A practical guide. Frontiers in Psychology, 4(AUG), 513. https://doi.org/10.3389/fpsyg.2013.00513

Labib, S. M., Lindley, S., & Huck, J. J. (2021). Estimating multiple greenspace exposure types and their associations with neighbourhood premature mortality: A socioecological study. Science of the Total Environment, 789, 147919. https://doi.org/10.1016/j.scitotenv.2021.147919

Lawlor, D. A., Harbord, R. M., Sterne, J. A. C., Timpson, N., & Davey Smith, G. (2008). Mendelian randomization: Using genes as instruments for making causal inferences in epidemiology. Statistics in Medicine, 27(8), 1133–1163. https://doi.org/10.1002/sim.3034

Li, D., Deal, B., Zhou, X., Slavenas, M., & Sullivan, W. C. (2018). Moving beyond the neighborhood: Daily exposure to nature and adolescents’ mood. Landscape and Urban Planning, 173, 33–43. https://doi.org/10.1016/j.landurbplan.2018.01.009

Manichaikul, A., Mychaleckyj, J. C., Rich, S. S., Daly, K., Sale, M., & Chen, W. M. (2010). Robust relationship inference in genome-wide association studies. Bioinformatics, 26(22), 2867–2873. https://doi.org/10.1093/bioinformatics/btq559

March, D., Hatch, S. L., Morgan, C., Kirkbride, J. B., Bresnahan, M., Fearon, P., & Susser, E. (2008). Psychosis and place. In Epidemiologic Reviews (Vol. 30, Issue 1, pp. 84–100). Oxford Academic. https://doi.org/10.1093/epirev/mxn006

McManus, S., Bebbington, P., Jenkins, R., Brugha, T., NHS Digital., & UK Statistics Authority. (2016). Mental health and wellbeing in England : Adult Psychiatric Morbidity Survey 2014.

Merlo, J., Chaix, B., Yang, M., Lynch, J., & Råstam, L. (2005). A brief conceptual tutorial of multilevel analysis in social epidemiology: Linking the statistical concept of clustering to the idea of contextual phenomenon. Journal of Epidemiology and Community Health, 59(6), 443–449. https://doi.org/10.1136/jech.2004.023473

Mitchell, R., Hemani, G., Dudding, T., Corbin, L., Harrison, S., & Paternoster, L. (2019). UK Biobank Genetic Data: MRC-IEU Quality Control, Version 2. https://doi.org/10.5523/bris.1ovaau5sxunp2cv8rcy88688v

Moore, T. H. M., Kesten, J. M., López-López, J. A., Ijaz, S., McAleenan, A., Richards, A., Gray, S., Savović, J., & Audrey, S. (2018). The effects of changes to the built environment on the mental health and well-being of adults: Systematic review. In Health and Place (Vol. 53, pp. 237–257). Pergamon. https://doi.org/10.1016/j.healthplace.2018.07.012

Morris, T. T., Davies, N. M., Hemani, G., & Smith, G. D. (2020). Population phenomena inflate genetic associations of complex social traits. Science Advances, 6(16), eaay0328. https://doi.org/10.1126/sciadv.aay0328

Mueller, M. A. E., Flouri, E., & Kokosi, T. (2019). The role of the physical environment in adolescent mental health. Health and Place, 58, 102153. https://doi.org/10.1016/j.healthplace.2019.102153

Office for National Statistics. (2020). 2011 *Census: Aggregate Data*. UK Data Service. http://doi.org/10.5257/census/aggregate-2011-2

Okbay, A., Baselmans, B. M. L., De Neve, J. E., Turley, P., Nivard, M. G., Fontana, M. A., Meddens, S. F. W., Linnér, R. K., Rietveld, C. A., Derringer, J., Gratten, J., Lee, J. J., Liu, J. Z., De Vlaming, R., SAhluwalia, T., Buchwald, J., Cavadino, A., Frazier-Wood, A. C., Furlotte, N. A., … Cesarini, D. (2016). Genetic variants associated with subjective well-being, depressive symptoms, and neuroticism identified through genome-wide analyses. Nature Genetics, 48(6), 624–633. https://doi.org/10.1038/ng.3552

Pedersen, C. B. (2015). Persons with schizophrenia migrate towards urban areas due to the development of their disorder or its prodromata. Schizophrenia Research, 168(1), 204–208. https://doi.org/10.1016/j.schres.2015.08.028

Peen, J., Schoevers, R. A., Beekman, A. T., & Dekker, J. (2010). The current status of urban-rural differences in psychiatric disorders. Acta Psychiatrica Scandinavica, 121(2), 84–93. https://doi.org/10.1111/j.1600-0447.2009.01438.x

Purcell, S., Neale, B., Todd-Brown, K., Thomas, L., Ferreira, M. A. R., Bender, D., Maller, J., Sklar, P., de Bakker, P. I. W., Daly, M. J., & Sham, P. C. (2007). PLINK: A Tool Set for Whole-Genome Association and Population-Based Linkage Analyses. The American Journal of Human Genetics, 81(3), 559–575. https://doi.org/10.1086/519795

Purtle, J., Nelson, K. L., Yang, Y., Langellier, B., Stankov, I., & Diez Roux, A. V. (2019). Urban–Rural Differences in Older Adult Depression: A Systematic Review and Meta-analysis of Comparative Studies. American Journal of Preventive Medicine, 56(4), 603–613. https://doi.org/10.1016/j.amepre.2018.11.008

R Core Team. (2016). R: A language and environment for statistical computing. R Foundation for Statistical Computing, Vienna, Austria.

Rhew, I. C., Vander Stoep, A., Kearney, A., Smith, N. L., & Dunbar, M. D. (2011). Validation of the Normalized Difference Vegetation Index as a Measure of Neighborhood Greenness. Annals of Epidemiology, 21(12), 946–952. https://doi.org/10.1016/j.annepidem.2011.09.001

Richmond, R. C., & Davey Smith, G. (2021). Mendelian Randomization: Concepts and Scope. *Cold Spring Harbor Perspectives in Medicine*, a040501. https://doi.org/10.1101/cshperspect.a040501

Ripke, S., Neale, B. M., Corvin, A., Walters, J. T. R., Farh, K.-H., Holmans, P. A., Lee, P., Bulik-Sullivan, B., Collier, D. A., Huang, H., Pers, T. H., Agartz, I., Agerbo, E., Albus, M., Alexander, M., Amin, F., Bacanu, S. A., Begemann, M., Belliveau Jr, R. A., … O’Donovan, M. C. (2014). Biological insights from 108 schizophrenia-associated genetic loci. Nature, 511(7510), 421–427. https://doi.org/10.1038/nature13595

Sariaslan, A., Fazel, S., D’Onofrio, B. M., Långström, N., Larsson, H., Bergen, S. E., Kuja-Halkola, R., & Lichtenstein, P. (2016). Schizophrenia and subsequent neighborhood deprivation: Revisiting the social drift hypothesis using population, twin and molecular genetic data. Translational Psychiatry, 6(5), e796–e796. https://doi.org/10.1038/tp.2016.62

Sarkar, C., Webster, C., & Gallacher, J. (2015). UK Biobank Urban Morphometric Platform (UKBUMP) – a nationwide resource for evidence-based healthy city planning and public health interventions. Annals of GIS, 21(2), 135–148. https://doi.org/10.1080/19475683.2015.1027791

Sarkar, C., Webster, C., & Gallacher, J. (2018). Residential greenness and prevalence of major depressive disorders: a cross-sectional, observational, associational study of 94 879 adult UK Biobank participants. The Lancet Planetary Health, 2(4), e162–e173. https://doi.org/10.1016/S2542-5196(18)30051-2

Sarmanova, A., Morris, T., & Lawson, D. J. (2020). Population stratification in GWAS meta-analysis should be standardized to the best available reference datasets. BioRxiv. https://doi.org/10.1101/2020.09.03.281568

Smith, D. J., Nicholl, B. I., Cullen, B., Martin, D., Ul-Haq, Z., Evans, J., Gill, J. M. R., Roberts, B., Gallacher, J., Mackay, D., Hotopf, M., Deary, I., Craddock, N., & Pell, J. P. (2013). Prevalence and characteristics of probable major depression and bipolar disorder within UK Biobank: Cross-sectional study of 172,751 participants. PLoS ONE, 8(11), e75362. https://doi.org/10.1371/journal.pone.0075362

Solmi, F., Dykxhoorn, J., & Kirkbride, J. B. (2017). Urban-Rural Differences in Major Mental Health Conditions (pp. 27–132). Springer, Singapore. https://doi.org/10.1007/978-981-10-2327-9_7

Sudlow, C., Gallacher, J., Allen, N., Beral, V., Burton, P., Danesh, J., Downey, P., Elliott, P., Green, J., Landray, M., Liu, B., Matthews, P., Ong, G., Pell, J., Silman, A., Young, A., Sprosen, T., Peakman, T., & Collins, R. (2015). UK Biobank: An Open Access Resource for Identifying the Causes of a Wide Range of Complex Diseases of Middle and Old Age. PLoS Medicine, 12(3), e1001779. https://doi.org/10.1371/journal.pmed.1001779

Tyrrell, J., Zheng, J., Beaumont, R., Hinton, K., Richardson, T. G., Wood, A. R., Davey Smith, G., Frayling, T. M., & Tilling, K. (2021). Genetic predictors of participation in optional components of UK Biobank. Nature Communications, 12(1), 886. https://doi.org/10.1038/s41467-021-21073-y

Vassos, E., Agerbo, E., Mors, O., & Bøcker Pedersen, C. (2016a). Urban-rural differences in incidence rates of psychiatric disorders in Denmark. British Journal of Psychiatry, 208(5), 435–440. https://doi.org/10.1192/bjp.bp.114.161091

Vassos, E., Agerbo, E., Mors, O., & Bøcker Pedersen, C. (2016b). Urban-rural differences in incidence rates of psychiatric disorders in Denmark. British Journal of Psychiatry, 208(5), 435–440. https://doi.org/10.1192/bjp.bp.114.161091

Verheij, R. A., Maas, J., & Groenewegen, P. P. (2008). Urban-rural health differences and the availability of green space. European Urban and Regional Studies, 15(4), 307–316. https://doi.org/10.1177/0969776408095107

Wang, P., Meng, Y. Y., Lam, V., & Ponce, N. (2019). Green space and serious psychological distress among adults and teens: A population-based study in California. Health and Place, 56, 184–190. https://doi.org/10.1016/j.healthplace.2019.02.002

Ward, J. S., Duncan, J. S., Jarden, A., & Stewart, T. (2016). The impact of children’s exposure to greenspace on physical activity, cognitive development, emotional wellbeing, and ability to appraise risk. Health and Place, 40, 44–50. https://doi.org/10.1016/j.healthplace.2016.04.015

Weeland, J., Laceulle, O. M., Nederhof, E., Overbeek, G., & Reijneveld, S. A. (2019). The greener the better? Does neighborhood greenness buffer the effects of stressful life events on externalizing behavior in late adolescence? Health and Place, 58, 102163. https://doi.org/10.1016/j.healthplace.2019.102163

White, M. P., Alcock, I., Wheeler, B. W., & Depledge, M. H. (2013). Would You Be Happier Living in a Greener Urban Area? A Fixed-Effects Analysis of Panel Data. Psychological Science, 24(6), 920–928. https://doi.org/10.1177/0956797612464659

Wray, N. R., Ripke, S., Mattheisen, M., Trzaskowski, M., Byrne, E. M., Abdellaoui, A., Adams, M. J., Agerbo, E., Air, T. M., Andlauer, T. M. F., Bacanu, S. A., Bækvad-Hansen, M., Beekman, A. F. T., Bigdeli, T. B., Binder, E. B., Blackwood, D. R. H., Bryois, J., Buttenschøn, H. N., Bybjerg-Grauholm, J., … Sullivan, P. F. (2018). Genome-wide association analyses identify 44 risk variants and refine the genetic architecture of major depression. Nature Genetics, 50(5), 668–681. https://doi.org/10.1038/s41588-018-0090-3

Zhang, Y., Mavoa, S., Zhao, J., Raphael, D., & Smith, M. (2020). The association between green space and adolescents mental well-being: A systematic review. International Journal of Environmental Research and Public Health, 17(18), 1–26. https://doi.org/10.3390/ijerph17186640

