## Supplementary materials for "Examining the association between genetic risk for depression, wellbeing and schizophrenia, and proximity to greenspace"

*1. Multilevel models (MLM)*

In our study we fit multilevel (or hierarchical/mixed) models (MLM) to estimate within-area effects of the polygenic score (PGS) exposure, whilst accounting for between-area differences in both the outcome (greenspace) and PGS exposure. A simplified specification is given in Equation 1:

$$Y_{ij}= \beta_{0j}+\beta_{1}x_{1ij}+\Sigma_{1}^{M}\beta_{M}x_{Mij}+u_{0j}+e_{0ij}$$

$$\beta_{0j}= \beta_{0}+u_{0j}$$

$$\left[ u_{0j} \right]\sim N\left( 0, \sigma_{u0}^{2} \right)$$

$$\left[ e_{0ij} \right]\sim N\left( 0, \sigma_{e0}^{2} \right)$$

Here $Y_{ij}$ denotes the greenspace outcome for individual *i* in neighbourhood *j*. Similarly, $x_{1ij}$ denotes the mental health PGS for individual *i* in neighbourhood *j,* and ${\Sigma_{1}^{M}x}_{Mij}$ denotes measures of individual covariates for the same individual *i* in neighbourhood *j.* $u_{0j}$ denotes the error term for each neighbourhood *j* around the global mean, while $e_{0ij}$ denotes the individual error term from the neighbourhood mean.

The multilevel specification allows us to model the greenspace outcome for individuals in a given neighbourhood, relative to the average greenspace in that neighbourhood. This is because the intercept term for neighbourhood *j*, $\beta_{0j}$, is constituted of two estimated parameters. $\beta_{0}$ gives the overall intercept, the average greenspace value for a typical individual in a typical neighbourhood, where all other covariates are zero.

Specific neighbourhood differentials are given by $u_{0j}$, this represents the difference in greenspace associated with living in neighbourhood *j*. These neighbourhood differentials are assumed to come from a normal distribution, with mean 0 and variance $\sigma_{u0}^{2}$. This variance represents the variation in greenspace outcomes between neighbourhoods.

$\beta_{1}$ then estimates the predicted greenspace change for a unit increase in mental health PGS, within the average neighbourhood, having taken account of between-neighbourhood differences via the random-intercept specification. $\beta_{M}$ represents the coefficients for remaining individual-level covariates in the adjustment set.

Finally, the residual unexplained, individual-level variation is given by $e_{0ij}$, and is similarly assumed to come from a normal distribution with mean 0 and variance $\sigma_{e0}^{2}$. From the variance components, $\sigma_{u0}^{2}$ and $\sigma_{e0}^{2}$, we can calculate the Variance Partitioning Coefficient (VPC), which assesses the proportion of total variation in outcome which is accounted for by a specific level in the model. In the two-level model formulation, the VPC is exactly equal to the intra-class correlation coefficient (ICC). The ICC gives the expected correlation between randomly selected pairs of lower level units from within the same higher level unit. For instance, we could calculate the Level-2 VPC for greenspace as follows:

$$Level 2 VPC for Y_{\mathrm{ij}}=\frac{\sigma_{u0}^{2}}{\sigma_{u0}^{2}+\sigma_{e0}^{2}}$$

In our example, this would tell us what proportion of the total unexplained variation in greenspace is accounted for by the neighbourhood in which someone lives. Similarly, in the ICC interpretation, it tells us the expected correlation between greenspace outcomes for pairs of individuals drawn from the same neighbourhood. Higher VPC or ICC values imply greater similarity in greenspace amongst individuals living in the same neighbourhood. This is particularly important for our investigation, as we know that greenspace exposure is likely to be very similar amongst individuals who live within the same neighbourhood – as it is not truly an individual level trait.

Supplementary Figure S1. R-squared values for models including increasing numbers of principal components for Normalised Difference Vegetation Index and percentage greenspace

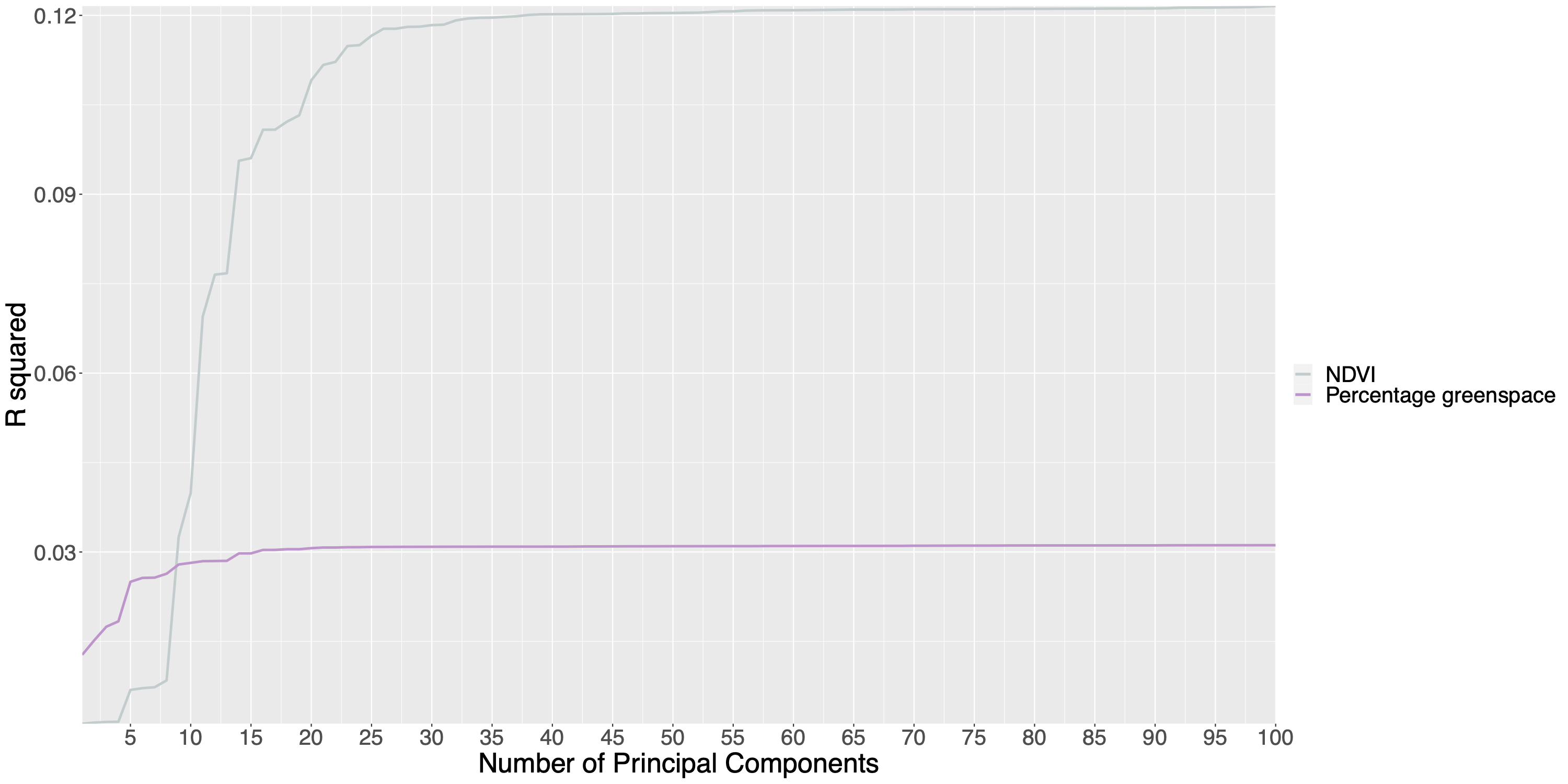

*NDVI= Normalised Difference Vegetation Index*

Supplementary Figure S2. Map of number of participants per neighbourhood in UK Biobank

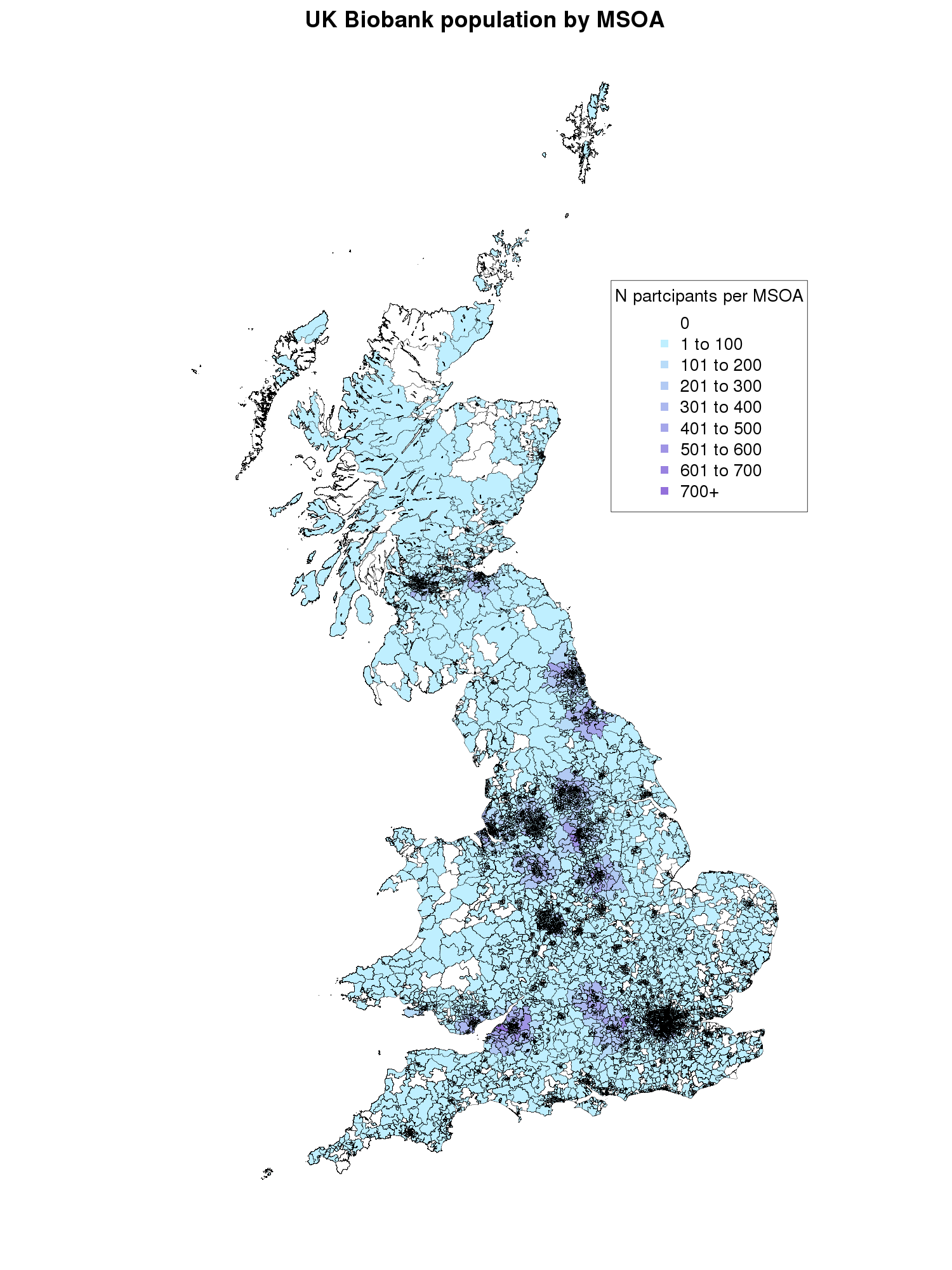

*Neighbourhoods refer to a combination of Middle Layer Super Output Areas (MSOAs) for England and Wales and Intermediate Zones (IZs) for Scotland*

Supplementary Figure S3. Map of number of participants per district in UK Biobank

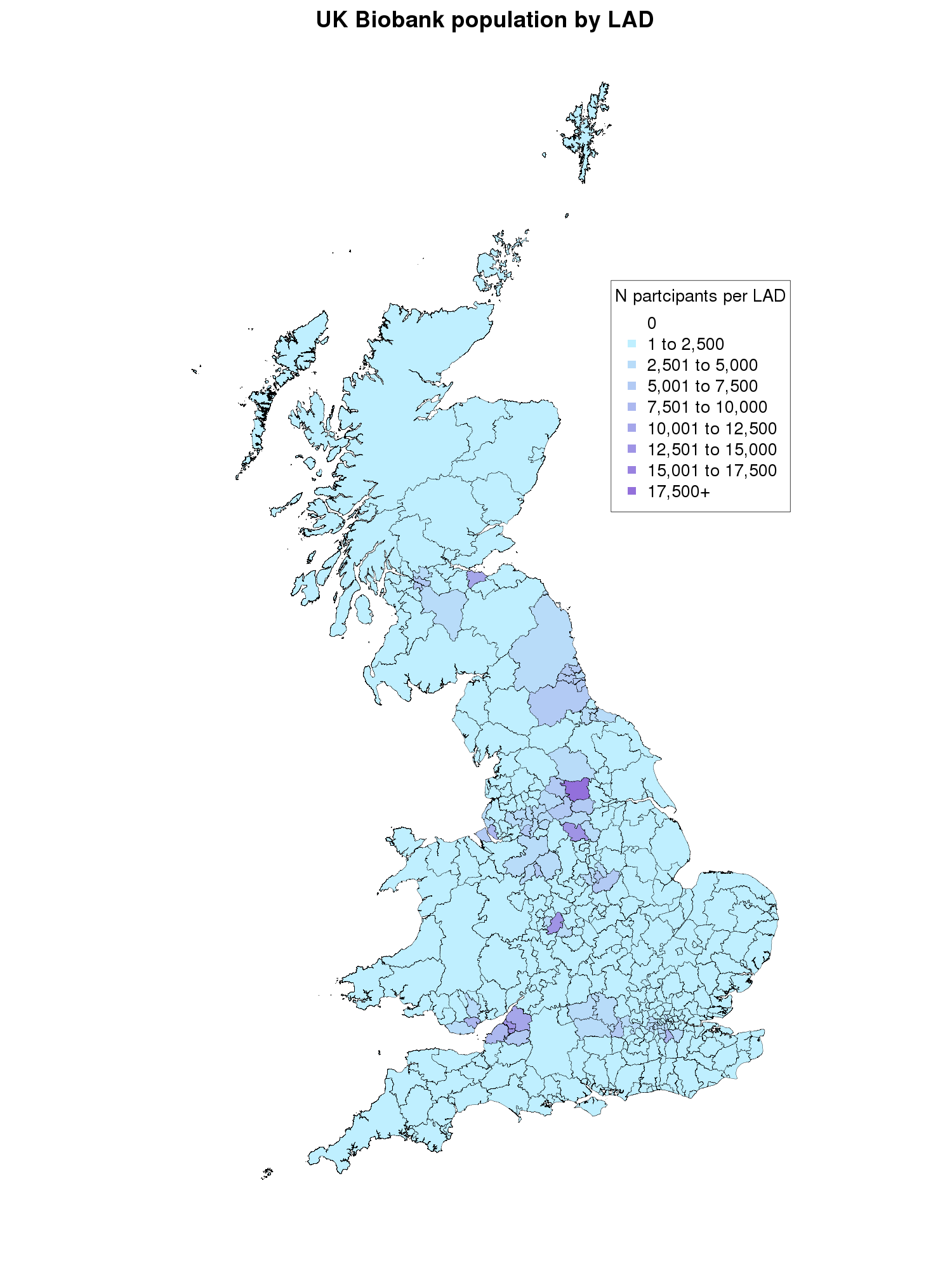

*Districts refer to a combination of Local Authority Districts (LADs) in England and Wales and Local Authorities (LAs) in Scotland*

Supplementary Table S1. Associations between depression, wellbeing and schizophrenia phenotypes and Normalised Difference Vegetation Index and percentage greenspace

| Mental health phenotype | Greenspace measure | N | Beta* | 2.5% CI | 97.5% CI | P |
| --- | --- | --- | --- | --- | --- | --- |
| *Depression diagnosis* | NDVI | 238,306 | -0.01 | -0.03 | -0.003 | 0.02 |
|  | Percentage greenspace | 293,922 | -0.06 | -0.07 | -0.05 | 6.04x10^-29^ |
| *MHQ depression* | NDVI | 64,740 | -0.01 | -0.03 | 0.0006 | 0.06 |
|  | Percentage greenspace | 77,358 | -0.03 | -0.05 | 0.02 | 1.52x10^-04^ |
| *Wellbeing* | NDVI | 76,443 | 0.008 | -0.001 | 0.02 | 0.09 |
|  | Percentage greenspace | 97,099 | 0.06 | 0.05 | 0.07 | 1.62x10^-41^ |
| *Schizophrenia* | NDVI | 238,306 | -0.03 | -0.13 | 0.07 | 0.53 |
|  | Percentage greenspace | 293,922 | -0.32 | -0.41 | -0.24 | 2.95x10^-13^ |

*CI=Confidence Interval, NDVI=Normalised Difference Vegetation Index, MHQ=Mental Health Questionnaire. *Beta reflects the NDVI or percentage greenspace SD change predicted by depression or schizophrenia diagnosis or every unit increase in the wellbeing score*

Supplementary Table S2. Associations between polygenic scores for depression, wellbeing and schizophrenia and Normalised Difference Vegetation Index for all p-value thresholds (N=238,306)

| Phenotype | Polygenic score p-value threshold | Beta* | 2.5% CI | 97.5% CI | P |
| --- | --- | --- | --- | --- | --- |
| *Depression* | 5x10^-08^ | 0.0002 | -0.004 | 0.004 | 0.92 |
|  | 1x10^-06^ | -0.002 | -0.005 | 0.002 | 0.43 |
|  | 1x10^-05^ | -0.002 | -0.006 | 0.002 | 0.27 |
|  | 1x10^-04^ | -0.003 | -0.007 | 0.001 | 0.15 |
|  | 1x10^-03^ | -0.005 | -0.009 | -0.001 | 0.01 |
|  | 0.01 | -0.007 | -0.01 | -0.003 | 0.0003 |
|  | 0.05 | -0.008 | -0.01 | -0.004 | 0.0001 |
|  | 0.1 | -0.008 | -0.01 | -0.004 | 0.00002 |
|  | 0.2 | -0.01 | -0.01 | -0.006 | 0.000001 |
|  | 0.3 | -0.01 | -0.01 | -0.006 | 0.000001 |
|  | 0.4 | -0.01 | -0.01 | -0.006 | 0.000001 |
|  | 0.5 | -0.01 | -0.01 | -0.006 | 0.000001 |
| *Wellbeing* | 1x10^-06^ | 0.003 | -0.001 | 0.007 | 0.13 |
|  | 1x10^-05^ | 0.003 | -0.001 | 0.006 | 0.17 |
|  | 1x10^-04^ | 0.003 | -0.0005 | 0.007 | 0.09 |
|  | 1x10^-03^ | -0.0006 | -0.0043 | 0.003 | 0.77 |
|  | 0.01 | -0.0005 | -0.004 | 0.003 | 0.81 |
|  | 0.05 | -0.0004 | -0.004 | 0.003 | 0.83 |
|  | 0.1 | 0.001 | -0.003 | 0.005 | 0.64 |
|  | 0.2 | 0.002 | -0.002 | 0.006 | 0.28 |
|  | 0.3 | 0.002 | -0.001 | 0.006 | 0.21 |
|  | 0.4 | 0.002 | -0.002 | 0.005 | 0.39 |
|  | 0.5 | 0.002 | -0.002 | 0.005 | 0.43 |
| *Schizophrenia* | 5x10^-08^ | 0.002 | -0.002 | 0.006 | 0.33 |
|  | 1x10^-06^ | 0.003 | -0.001 | 0.006 | 0.15 |
|  | 1x10^-05^ | 0.002 | -0.001 | 0.006 | 0.22 |
|  | 1x10^-04^ | 0.001 | -0.003 | 0.005 | 0.52 |
|  | 1x10^-03^ | 0.002 | -0.002 | 0.006 | 0.32 |
|  | 0.01 | 0.002 | -0.002 | 0.006 | 0.35 |
|  | 0.05 | 0.0005 | -0.004 | 0.005 | 0.80 |
|  | 0.1 | 0.0004 | -0.004 | 0.004 | 0.86 |
|  | 0.2 | 0.0004 | -0.004 | 0.004 | 0.86 |
|  | 0.3 | -0.0002 | -0.004 | 0.004 | 0.92 |
|  | 0.4 | -0.0004 | -0.004 | 0.004 | 0.85 |
|  | 0.5 | -0.0002 | -0.004 | 0.004 | 0.93 |

*CI=Confidence Interval. *Beta reflects the NDVI SD change predicted by an SD increase in the PGS*

Supplementary Table S3. Associations between polygenic scores for depression, wellbeing and schizophrenia and percentage greenspace for all p-value thresholds (N=293,922)

| Phenotype | Polygenic score p-value threshold | Beta | 2.5% CI | 97.5% CI | P |
| --- | --- | --- | --- | --- | --- |
| *Depression* | 5x10^-08^ | -0.004 | -0.007 | -0.0003 | 0.03 |
|  | 1x10^-06^ | 0.001 | -0.002 | 0.005 | 0.50 |
|  | 1x10^-05^ | 0.00003 | -0.004 | 0.004 | 0.99 |
|  | 1x10^-04^ | -0.0004 | -0.004 | 0.003 | 0.82 |
|  | 1x10^-03^ | -0.003 | -0.006 | 0.0008 | 0.12 |
|  | 0.01 | -0.005 | -0.009 | -0.001 | 0.006 |
|  | 0.05 | -0.005 | -0.009 | -0.001 | 0.007 |
|  | 0.1 | -0.005 | -0.009 | -0.002 | 0.004 |
|  | 0.2 | -0.006 | -0.01 | -0.003 | 0.0005 |
|  | 0.3 | -0.007 | -0.01 | -0.003 | 0.0005 |
|  | 0.4 | -0.006 | -0.01 | -0.003 | 0.0007 |
|  | 0.5 | -0.006 | -0.01 | -0.003 | 0.0007 |
| *Wellbeing* | 1x10^-06^ | 0.001 | -0.003 | 0.005 | 0.58 |
|  | 1x10^-05^ | 0.003 | -0.0009 | 0.006 | 0.14 |
|  | 1x10^-04^ | 0.002 | -0.002 | 0.005 | 0.37 |
|  | 1x10^-03^ | 0.003 | -0.0005 | 0.007 | 0.09 |
|  | 0.01 | 0.006 | 0.003 | 0.01 | 0.0006 |
|  | 0.05 | 0.006 | 0.003 | 0.01 | 0.0005 |
|  | 0.1 | 0.007 | 0.003 | 0.01 | 0.0003 |
|  | 0.2 | 0.007 | 0.003 | 0.01 | 0.0002 |
|  | 0.3 | 0.007 | 0.003 | 0.01 | 0.0002 |
|  | 0.4 | 0.006 | 0.003 | 0.01 | 0.0009 |
|  | 0.5 | 0.006 | 0.002 | 0.01 | 0.001 |
| *Schizophrenia* | 5x10^-08^ | 0.002 | -0.002 | 0.005 | 0.35 |
|  | 1x10^-06^ | 0.001 | -0.002 | 0.005 | 0.49 |
|  | 1x10^-05^ | 0.0004 | -0.003 | 0.004 | 0.83 |
|  | 1x10^-04^ | 0.00009 | -0.004 | 0.004 | 0.96 |
|  | 1x10^-03^ | -0.002 | -0.006 | 0.002 | 0.31 |
|  | 0.01 | -0.003 | -0.007 | 0.0005 | 0.09 |
|  | 0.05 | -0.004 | -0.008 | -0.0006 | 0.02 |
|  | 0.1 | -0.005 | -0.009 | -0.0009 | 0.02 |
|  | 0.2 | -0.005 | -0.009 | -0.0009 | 0.02 |
|  | 0.3 | -0.004 | -0.008 | 0.0003 | 0.07 |
|  | 0.4 | -0.004 | -0.008 | -0.0005 | 0.03 |
|  | 0.5 | -0.005 | -0.009 | -0.0008 | 0.02 |

*CI=Confidence Interval. *Beta reflects the percentage greenspace SD change predicted by an SD increase in the PGS*

Supplementary Table S4. Akaike’s Information Criteria (AIC) for linear regression model and the neighbourhood and district area multilevel models

|  |  | AIC for percentage greenspace | | | AIC for NDVI measure | | |
| --- | --- | --- | --- | --- | --- | --- | --- |
| Phenotype | Polygenic score p-value threshold | Linear model | Neighbourhood model | District model | Linear model | Neighbourhood model | District model |
| *Depression* | 5x10^-08^ | 831302.43 | 645828.05 | 749931.02 | 644899.32 | 228996.94 | 345494.96 |
|  | 1x10^-06^ | 831306.47 | 645829.75 | 749934.98 | 644898.71 | 228996.79 | 345494.97 |
|  | 1x10^-05^ | 831306.91 | 645830.25 | 749934.79 | 644898.13 | 228995.28 | 345494.20 |
|  | 1x10^-04^ | 831306.86 | 645830.42 | 749935.10 | 644897.24 | 228992.37 | 345493.14 |
|  | 1x10^-03^ | 831304.54 | 645830.13 | 749932.17 | 644893.20 | 228996.87 | 345492.26 |
|  | 0.01 | 831299.35 | 645827.37 | 749921.85 | 644886.54 | 228995.88 | 345489.43 |
|  | 0.05 | 831299.65 | 645822.73 | 749916.49 | 644883.63 | 228990.92 | 345481.63 |
|  | 0.1 | 831298.64 | 645823.15 | 749917.33 | 644881.02 | 228989.60 | 345481.47 |
|  | 0.2 | 831294.72 | 645817.52 | 749909.81 | 644874.54 | 228987.08 | 345481.20 |
|  | 0.3 | 831294.67 | 645817.61 | 749909.82 | 644874.76 | 228987.77 | 345480.94 |
|  | 0.4 | 831295.38 | 645817.41 | 749910.20 | 644874.52 | 228988.82 | 345481.72 |
|  | 0.5 | 831295.46 | 645818.11 | 749909.72 | 644874.41 | 228989.37 | 345482.66 |
| *Wellbeing* | 1x10^-06^ | 831306.61 | 645829.45 | 749934.59 | 644897.04 | 228991.58 | 345490.05 |
|  | 1x10^-05^ | 831304.78 | 645830.00 | 749934.32 | 644897.44 | 228996.08 | 345493.98 |
|  | 1x10^-04^ | 831306.12 | 645830.51 | 749934.35 | 644896.39 | 228995.80 | 345491.67 |
|  | 1x10^-03^ | 831304.00 | 645828.80 | 749931.30 | 644899.24 | 228996.38 | 345493.36 |
|  | 0.01 | 831295.25 | 645826.92 | 749927.56 | 644899.27 | 228995.40 | 345493.32 |
|  | 0.05 | 831294.61 | 645828.12 | 749928.85 | 644899.28 | 228995.35 | 345492.78 |
|  | 0.1 | 831293.63 | 645825.17 | 749926.12 | 644899.11 | 228994.51 | 345493.35 |
|  | 0.2 | 831292.91 | 645823.00 | 749922.06 | 644898.14 | 228994.52 | 345493.24 |
|  | 0.3 | 831293.26 | 645823.91 | 749922.75 | 644897.73 | 228994.45 | 345492.93 |
|  | 0.4 | 831295.83 | 645824.86 | 749925.03 | 644898.60 | 228995.49 | 345493.53 |
|  | 0.5 | 831296.14 | 645824.63 | 749925.72 | 644898.70 | 228995.46 | 345493.81 |
| *Schizophrenia* | 5x10^-08^ | 831306.03 | 645826.85 | 749932.54 | 644898.37 | 228996.43 | 345493.87 |
|  | 1x10^-06^ | 831306.44 | 645826.34 | 749932.07 | 644897.29 | 228995.53 | 345493.39 |
|  | 1x10^-05^ | 831306.87 | 645825.87 | 749932.61 | 644897.85 | 228995.47 | 345493.57 |
|  | 1x10^-04^ | 831306.91 | 645825.74 | 749933.15 | 644898.91 | 228996.36 | 345494.67 |
|  | 1x10^-03^ | 831305.89 | 645823.87 | 749932.82 | 644898.33 | 228996.47 | 345494.63 |
|  | 0.01 | 831304.09 | 645825.32 | 749932.25 | 644898.45 | 228996.82 | 345494.87 |
|  | 0.05 | 831301.80 | 645825.88 | 749934.51 | 644899.26 | 228996.19 | 345494.45 |
|  | 0.1 | 831301.19 | 645827.30 | 749934.86 | 644899.30 | 228996.56 | 345494.65 |
|  | 0.2 | 831301.19 | 645827.30 | 749934.86 | 644899.30 | 228996.56 | 345494.65 |
|  | 0.3 | 831303.62 | 645825.02 | 749934.47 | 644899.32 | 228996.73 | 345494.76 |
|  | 0.4 | 831302.02 | 645826.41 | 749934.84 | 644899.29 | 228996.75 | 345494.29 |
|  | 0.5 | 831301.25 | 645826.98 | 749934.92 | 644899.32 | 228996.76 | 345494.14 |

*AIC=* *Akaike’s Information Criteria, Neighbourhood= Middle Layer Super Output Area or intermediate zone, District= Local Authority District or Local Authority, NDVI= Normalised Difference Vegetation Index*

Supplementary Table S5. Associations between polygenic scores for depression, wellbeing and schizophrenia and Normalised Difference Vegetation Index whilst allowing for different intercepts for each area (neighbourhood and district) for all p-value thresholds (N=238,306)

|  |  | Neighbourhood | | | | District | | | |
| --- | --- | --- | --- | --- | --- | --- | --- | --- | --- |
| Phenotype | **Polygenic score p-value threshold** | **Beta*** | **2.5% CI** | **97.5% CI** | **P** | **Beta*** | **2.5% CI** | **97.5% CI** | **P** |
| *Depression* | 5x10^-08^ | 0.0002 | -0.001 | 0.002 | 0.83 | 0.0001 | -0.002 | 0.002 | 0.90 |
|  | 1x10^-06^ | -0.0003 | -0.002 | 0.001 | 0.70 | -0.0001 | -0.002 | 0.002 | 0.89 |
|  | 1x10^-05^ | 0.001 | -0.0005 | 0.003 | 0.19 | 0.0009 | -0.001 | 0.003 | 0.37 |
|  | 1x10^-04^ | 0.002 | 0.0001 | 0.003 | 0.03 | 0.001 | -0.0007 | 0.003 | 0.19 |
|  | 1x10^-03^ | 0.0003 | -0.001 | 0.002 | 0.75 | -0.002 | -0.004 | 0.0003 | 0.10 |
|  | 0.01 | -0.0008 | -0.002 | 0.0008 | 0.33 | -0.002 | -0.004 | -0.0005 | 0.02 |
|  | 0.05 | -0.002 | -0.003 | -0.0003 | 0.02 | -0.004 | -0.006 | -0.002 | 0.0002 |
|  | 0.1 | -0.002 | -0.004 | -0.0005 | 0.01 | -0.004 | -0.006 | -0.002 | 0.0002 |
|  | 0.2 | -0.002 | -0.004 | -0.0008 | 0.002 | -0.004 | -0.006 | -0.002 | 0.0001 |
|  | 0.3 | -0.002 | -0.004 | -0.0008 | 0.003 | -0.004 | -0.006 | -0.002 | 0.0001 |
|  | 0.4 | -0.002 | -0.004 | -0.0006 | 0.006 | -0.004 | -0.006 | -0.002 | 0.0002 |
|  | 0.5 | -0.002 | -0.004 | -0.0005 | 0.009 | -0.004 | -0.006 | -0.002 | 0.0003 |
| *Wellbeing* | 1x10^-06^ | 0.002 | 0.0003 | 0.003 | 0.02 | 0.002 | 0.0003 | 0.004 | 0.03 |
|  | 1x10^-05^ | 0.0008 | -0.0007 | 0.002 | 0.30 | 0.001 | -0.0009 | 0.003 | 0.28 |
|  | 1x10^-04^ | 0.0008 | -0.0007 | 0.002 | 0.29 | 0.002 | -0.00002 | 0.004 | 0.05 |
|  | 1x10^-03^ | 0.0006 | -0.0009 | 0.002 | 0.43 | 0.001 | -0.0006 | 0.003 | 0.18 |
|  | 0.01 | 0.001 | -0.0006 | 0.002 | 0.22 | 0.001 | -0.0006 | 0.003 | 0.16 |
|  | 0.05 | 0.001 | -0.0006 | 0.003 | 0.22 | 0.002 | -0.0004 | 0.004 | 0.11 |
|  | 0.1 | 0.001 | -0.0004 | 0.003 | 0.14 | 0.001 | -0.0006 | 0.003 | 0.17 |
|  | 0.2 | 0.001 | -0.0004 | 0.003 | 0.14 | 0.001 | -0.0005 | 0.004 | 0.15 |
|  | 0.3 | 0.001 | -0.0003 | 0.003 | 0.12 | 0.002 | -0.0004 | 0.004 | 0.12 |
|  | 0.4 | 0.0009 | -0.0006 | 0.002 | 0.24 | 0.001 | -0.0007 | 0.003 | 0.19 |
|  | 0.5 | 0.0009 | -0.0006 | 0.002 | 0.23 | 0.001 | -0.0008 | 0.003 | 0.23 |
| *Schizophrenia* | 5x10^-08^ | 0.0006 | -0.0009 | 0.002 | 0.42 | 0.001 | -0.0010 | 0.003 | 0.32 |
|  | 1x10^-06^ | 0.001 | -0.0005 | 0.002 | 0.21 | 0.001 | -0.0008 | 0.003 | 0.23 |
|  | 1x10^-05^ | 0.001 | -0.0005 | 0.003 | 0.20 | 0.001 | -0.0008 | 0.003 | 0.26 |
|  | 1x10^-04^ | 0.0007 | -0.0008 | 0.002 | 0.39 | 0.0005 | -0.002 | 0.002 | 0.63 |
|  | 1x10^-03^ | 0.0006 | -0.0009 | 0.002 | 0.44 | 0.0005 | -0.001 | 0.003 | 0.61 |
|  | 0.01 | -0.0002 | -0.002 | 0.001 | 0.85 | 0.0002 | -0.002 | 0.002 | 0.87 |
|  | 0.05 | -0.0006 | -0.002 | 0.001 | 0.45 | -0.0007 | -0.003 | 0.001 | 0.52 |
|  | 0.1 | -0.0004 | -0.002 | 0.001 | 0.66 | -0.0005 | -0.003 | 0.002 | 0.67 |
|  | 0.2 | -0.0004 | -0.002 | 0.001 | 0.66 | -0.0005 | -0.003 | 0.002 | 0.67 |
|  | 0.3 | -0.0002 | -0.002 | 0.002 | 0.85 | -0.0002 | -0.002 | 0.002 | 0.85 |
|  | 0.4 | -0.0001 | -0.002 | 0.002 | 0.87 | -0.0008 | -0.003 | 0.001 | 0.47 |
|  | 0.5 | -0.0001 | -0.002 | 0.002 | 0.87 | -0.0009 | -0.003 | 0.001 | 0.42 |

*CI=Confidence Interval, Neighbourhood= Middle Layer Super Output Area or intermediate zone, District= Local Authority District or Local Authority. *Beta reflects the NDVI SD change predicted by an SD increase in the PGS, for the average individual in a typical neighbourhood or district.*

Supplementary Table S6. Associations between polygenic scores for depression, wellbeing and schizophrenia and percentage greenspace, whilst allowing for different intercepts for each area (neighbourhood and district) for all p-value thresholds (N=293,922)

|  |  | Neighbourhood | | | | District | | | |
| --- | --- | --- | --- | --- | --- | --- | --- | --- | --- |
| Phenotype | **Polygenic score p-value threshold** | **Beta*** | **2.5% CI** | **97.5% CI** | **P** | **Beta*** | **2.5% CI** | **97.5% CI** | **P** |
| *Depression* | 5x10^-08^ | -0.002 | -0.005 | 0.0005 | 0.12 | -0.003 | -0.006 | -0.00008 | 0.04 |
|  | 1x10^-06^ | 0.001 | -0.001 | 0.004 | 0.36 | 0.0006 | -0.003 | 0.004 | 0.72 |
|  | 1x10^-05^ | 0.001 | -0.002 | 0.003 | 0.59 | 0.0009 | -0.002 | 0.004 | 0.58 |
|  | 1x10^-04^ | 0.0005 | -0.002 | 0.003 | 0.72 | 0.00008 | -0.003 | 0.003 | 0.96 |
|  | 1x10^-03^ | -0.0008 | -0.003 | 0.002 | 0.56 | -0.003 | -0.006 | 0.0003 | 0.08 |
|  | 0.01 | -0.002 | -0.005 | 0.0003 | 0.08 | -0.006 | -0.009 | -0.003 | 0.0002 |
|  | 0.05 | -0.004 | -0.006 | -0.001 | 0.006 | -0.007 | -0.01 | -0.004 | 0.00001 |
|  | 0.1 | -0.004 | -0.006 | -0.0009 | 0.008 | -0.007 | -0.01 | -0.004 | 0.00002 |
|  | 0.2 | -0.005 | -0.007 | -0.002 | 0.0004 | -0.008 | -0.01 | -0.005 | 0.0000003 |
|  | 0.3 | -0.005 | -0.007 | -0.002 | 0.0004 | -0.008 | -0.01 | -0.005 | 0.0000003 |
|  | 0.4 | -0.005 | -0.007 | -0.002 | 0.0003 | -0.008 | -0.01 | -0.005 | 0.0000004 |
|  | 0.5 | -0.005 | -0.007 | -0.002 | 0.0005 | -0.008 | -0.01 | -0.005 | 0.0000003 |
| *Wellbeing* | 1x10^-06^ | 0.001 | -0.001 | 0.004 | 0.27 | 0.001 | -0.002 | 0.004 | 0.48 |
|  | 1x10^-05^ | 0.001 | -0.002 | 0.004 | 0.45 | 0.001 | -0.002 | 0.005 | 0.35 |
|  | 1x10^-04^ | 0.00002 | -0.003 | 0.003 | 0.99 | 0.001 | -0.002 | 0.005 | 0.37 |
|  | 1x10^-03^ | 0.002 | -0.0009 | 0.004 | 0.20 | 0.003 | -0.00001 | 0.006 | 0.05 |
|  | 0.01 | 0.002 | -0.0001 | 0.005 | 0.06 | 0.004 | 0.001 | 0.008 | 0.006 |
|  | 0.05 | 0.002 | -0.0006 | 0.005 | 0.14 | 0.004 | 0.0009 | 0.007 | 0.01 |
|  | 0.1 | 0.003 | 0.0003 | 0.006 | 0.03 | 0.005 | 0.002 | 0.008 | 0.003 |
|  | 0.2 | 0.003 | 0.0009 | 0.006 | 0.009 | 0.006 | 0.003 | 0.009 | 0.0003 |
|  | 0.3 | 0.003 | 0.0006 | 0.006 | 0.01 | 0.006 | 0.002 | 0.009 | 0.0005 |
|  | 0.4 | 0.003 | 0.0004 | 0.006 | 0.02 | 0.005 | 0.002 | 0.008 | 0.002 |
|  | 0.5 | 0.003 | 0.0005 | 0.006 | 0.02 | 0.005 | 0.002 | 0.008 | 0.002 |
| *Schizophrenia* | 5x10^-08^ | 0.003 | 0.00009 | 0.005 | 0.04 | 0.003 | -0.0006 | 0.006 | 0.11 |
|  | 1x10^-06^ | 0.003 | 0.0003 | 0.005 | 0.03 | 0.003 | -0.0004 | 0.006 | 0.08 |
|  | 1x10^-05^ | 0.003 | 0.0004 | 0.006 | 0.02 | 0.002 | -0.0006 | 0.006 | 0.12 |
|  | 1x10^-04^ | 0.003 | 0.0005 | 0.006 | 0.02 | 0.002 | -0.0009 | 0.005 | 0.17 |
|  | 1x10^-03^ | 0.004 | 0.001 | 0.006 | 0.007 | 0.002 | -0.0008 | 0.006 | 0.14 |
|  | 0.01 | 0.003 | 0.0005 | 0.006 | 0.02 | 0.003 | -0.0005 | 0.006 | 0.10 |
|  | 0.05 | 0.003 | 0.0003 | 0.006 | 0.03 | 0.001 | -0.002 | 0.004 | 0.51 |
|  | 0.1 | 0.003 | -0.0003 | 0.005 | 0.08 | 0.0005 | -0.003 | 0.004 | 0.79 |
|  | 0.2 | 0.003 | -0.0003 | 0.005 | 0.08 | 0.0005 | -0.003 | 0.004 | 0.79 |
|  | 0.3 | 0.003 | 0.0005 | 0.006 | 0.02 | 0.001 | -0.002 | 0.005 | 0.51 |
|  | 0.4 | 0.003 | 0.00006 | 0.006 | 0.05 | 0.0005 | -0.003 | 0.004 | 0.77 |
|  | 0.5 | 0.003 | -0.0002 | 0.005 | 0.06 | 0.0002 | -0.003 | 0.004 | 0.92 |

*CI=Confidence Interval, Neighbourhood= Middle Layer Super Output Area or intermediate zone, District= Local Authority District or Local Authority. *Beta reflects the percentage greenspace SD change predicted by an SD increase in the PGS, for the average individual in a typical neighbourhood or district.*

Supplementary Table S7. Sensitivity analyses for one-sample Mendelian randomisation

|  |  | IVW | | | MR Egger | | | LAD | | MR-RAPS | |
| --- | --- | --- | --- | --- | --- | --- | --- | --- | --- | --- | --- |
| Exposure | **Outcome** | **Beta* (95% CI)** | **P** | **Mean F statistic** | **Beta* (95% CI)** | **P** | **I-squared** | **Beta* (95% CI)** | **P** | **Beta* (95% CI)** | **P** |
| Depression  diagnosis | NDVI | 0.29  (-0.22, 0.80) | 0.26 | 8.18 | 0.79  (-1.08, 2.67) | 0.41 | 0.16 | 0.04  (-0.84, 0.93) | 0.92 | NA | NA |
| MHQ depression | NDVI | 0.08  (-0.43, 0.59) | 0.76 | 3.08 | -0.32  (-1.24, 0.60) | 0.50 | 0.05 | 0.02  (-0.50, 0.54) | 0.93 | NA | NA |
| Depression  diagnosis | Percentage greenspace | -0.22  (-0.74, 0.31) | 0.41 | 8.38 | 1.51  (-0.16, 3.17) | 0.08 | 0.27 | 0.21  (-0.77, 1.18) | 0.68 | NA | NA |
| MHQ depression | Percentage greenspace | -0.49  (-1.16, 0.18) | 0.15 | 3.93 | -0.59  (-1.95, 0.76) | 0.39 | 0.15 | 0.25  (-0.87, 1.37) | 0.66 | NA | NA |
| Wellbeing  (GRS at 1x10^-05^) | Percentage greenspace | -0.80  (-3.92, 2.31) | 0.61 | 1.22 | -0.86  (-2.33, 0.60) | 0.25 | 0.00 | 0.24  (-1.07, 1.54) | 0.72 | 1.26  (-0.28, 2.80) | 0.11 |
| Schizophrenia | Percentage greenspace | -5.06  (-22.37, 12.25) | 0.57 | 1.27 | -4.85  (-13.22, 3.52) | 0.26 | 0.00 | -2.76  (-12.87, 7.35) | 0.59 | NA | NA |

**Beta reflects the NDVI or percentage greenspace SD predicted by an increased genetic liability for depression and schizophrenia or every one unit increase in the wellbeing score*
